# Plasma proteome profiling identifies predictive signatures for preterm birth risk

**DOI:** 10.1101/2025.09.24.25336553

**Authors:** Naman Kharbanda, Ankit Biswas, Ramachandran Thiruvengadam, Shakuntala Bai, Surya Sankar Haldar, Arundhati Tiwari, Sandhini Saha, Nikhil Sharma, Bapu Koundinya Desiraju, Pallavi Kshetrapal, Nitya Wadhwa, Dinakar M Salunke, Shinjini Bhatnagar, Tushar Kanti Maiti, GARBH-Ini study group

**Affiliations:** Regional Centre for Biotechnology, NCR Biotech Science Cluster, Faridabad 121001, India; Translational Health Science and Technology Institute, NCR Biotech Science Cluster, Faridabad 121001, India; International Centre for Genetic Engineering and Biotechnology, Aruna Asaf Ali Marg, New Delhi, 110067, India

## Abstract

Preterm birth (PTB) is a major public health concern, and its associated complications account for 16.6% of deaths in children under five years. Over 50% of such deliveries are spontaneous, with unknown underlying causes. We identified significant changes in the plasma proteome across mid-trimester prior to the pre-term deliveries, indicating signals for identifying mothers at risk for PTB. Using quantitative LC-MS analysis and machine learning, we identified high plasma levels of Calcyophosin-2 (CAPS2) at 18-20 weeks of gestation in women who delivered preterm. Prediction model based on plasma CAPS2 level in a case-cohort (n=795) design accurately predicted high-risk sPTB with a detection rate of over 90% while reducing 40% false positivity and therefore avoiding subsequent unnecessary tests in them. Our findings strongly highlight CAPS2 as a novel candidate biomarker for prediction at early mid-trimester, accurately screening women at risk of preterm delivery, particularly in low-resource primary and secondary care settings where there are no existing screening programs for preterm birth.

**Teaser:** Mid-trimester screening of mothers at risk of delivering preterm babies using a single marker test.

## Introduction

Preterm birth (PTB) is defined as a live birth before 37 weeks of completed gestation^1^. It is one of the leading causes of newborn mortality and lifelong morbidities. Of the 5.3 million deaths reported for children under five years of age in 2019, ∼16.6% (approximately 0.94 million) were attributed to preterm birth-associated complications^2^. The surviving preterm babies are predisposed to both short-term and long-term challenges of physical, developmental, and socioeconomic nature^3^. The global prevalence is estimated to stand at 13.4 million preterm live births annually. India’s contribution to the global burden of PTB comes with 13% of the annual rate (3.02 million cases in 2020), the third highest after Bangladesh and Pakistan^4^. The vast majority are spontaneous (sPTB), where the reasons for premature birth remain unpredictable^5^.

Pregnancy is a highly intricate process marked by a series of dynamic and well-coordinated changes that enable the transformation of a single-cell zygote into a fully developed fetus^6^. The onset of labor is characterized by a series of coordinated physiological processes in the uterus, including cervical ripening, activation of the amniotic and decidual membranes, and a shift in the uterine smooth muscle from a relaxed to contractile state^7^. Thus, timely and safe parturition is the key to a successful pregnancy. Disturbances in this well-balanced machinery have the potential to translate into major adverse pregnancy outcomes. Plasma is routinely used to screen for different pregnancy-associated clinical parameters as the circulating biomolecules are a reliable reflection of the feto-maternal system. The risk of pregnancy complications has been assessed using electronic health records that include maternal characteristics, obstetric and gynaecological history, as well as the features of the current pregnancy^8^. Several studies have also highlighted potential protein biomarkers, such as Interleukin-6 (*IL6*), Serum amyloid A (*SAA1*), and Pregnancy-associated Plasma Protein A (*PAPPA*). However, these biomarkers have limitations due to small sample size, insubstantial predictability, lack of validation, and applicability^9,10^. Many of the potential markers, such as fetal fibronectin (*FN1*), or Placental Alpha Macroglobulin 1 (*PAMG-1*) are suitable for symptomatic pregnancies, i.e., in women with signs of preterm labor (PTL), hindering their clinical utility^11,12^. The ratio of Insulin-like Growth Factor Binding Protein 4 (*IGFBP4*) to Sex Hormone Binding Globulin (*SHBG*) in serum/plasma is one of the most extensively studied biomarkers for predicting asymptomatic spontaneous preterm birth (sPTB). *IBP4*/*SHBG* ratio was identified as a biomarker through LC-MS analysis of high-abundant protein-depleted serum. However, this pipeline identified only 147 proteins, potentially missing a large number of low-abundance serum proteins that may also serve as biomarkers for the sPTB^13^. In a nested case-control study of Asian and sub-Saharan cohorts, this marker achieved an AUROC of 0.55 for the prediction of sPTB.^14^. This underscores the need for advanced mass spectrometry-based discovery proteomics to identify low-abundance serum/plasma proteins and develop and validate a sensitive early-stage predictive or screening marker for preterm birth using clinically appropriate study designs.

Recent advancements in mass spectrometry-based plasma proteomics, particularly the data-independent acquisition (DIA) approach, such as SWATH-MS, have revolutionized biomarker discovery in pregnancy research. These techniques enable comprehensive profiling of the plasma proteome, even in neat plasma, offering new opportunities to identify novel biomarkers for PTB beyond the most abundant proteins. The objective of this study is to identify plasma proteome signatures across mid-trimester and develop an early-stage (at 18-20 weeks of pregnancy) and sensitive biomarker for accurately screening or predicting women at risk of sPTB, particularly the severe form where the baby is born ≤32 weeks of gestation. In phase 1 of the study, which consisted of discovery and verification arms, samples were selected in a case-control design (n=27) at two time points, viz., early mid-trimester (EMT, 18-20 weeks of gestation) and late mid-trimester (LMT, 26-28 weeks of gestation). Plasma samples were subjected to global proteomics analysis, in which we identified 113 and 90 differentially expressed proteins (DEPs) at EMT and LMT, respectively. Further downstream, targeted measurement of selected proteins and ML-based feature extraction filtered out *MICAL2* and *CAPS2* as important predictors of sPTB. In phase 2, these selected protein signatures were validated in a separate sample set in a case-cohort study design (n=795). Finally, the prediction models were developed utilizing the observations from proteome data for early-stage screening of sPTB as well as high-risk sPTB cases. Model 2, developed using the plasma levels of CAPS2, performed best, demonstrating a sensitivity of ∼90% (95% CI: 81-98%) and a specificity of 25.2% (95% CI: 22-28%). In high-risk sPTB cases, the model performance improved, achieving more than 90% (95% CI: 78-100%) sensitivity for a specificity of 39% (95% CI: 36-42%). High levels of *CAPS2*, a calcium-binding protein, in plasma can interfere with the intricate balance of Ca^2+^ in the body fluids and smooth muscles, which is crucial for maintaining a healthy pregnancy and a successful delivery^15–17^

## Results

### Clinical and socio-demographic characteristics of the participants at baseline and delivery

The study was conducted in two phases. In Phase 1, with the aim of identifying proteome-wide molecular signatures associated with sPTB, a nested case-control study was conducted within the GARBH-Ini cohort. The study included pregnant women with available plasma samples at both EMT (18–20 weeks) and LMT (26–28 weeks) timepoints. Participants who delivered before 35 weeks of gestation were classified as sPTB (case), while those who delivered at or beyond 39 weeks were considered TB (controls). Eventually, 27 matched case-control pairs were selected at each time point for the proteomic data acquisition and analysis (Fig. 1 and 2A). The median age of the selected mothers was 23 years (Inter quartile range (IQR): 21, 25) at the first antenatal care (ANC) visit, whereas the median body weight was 43 kg (IQR: 40, 53). About 50% of the women included in this study were nulliparous. No substantial differences were observed in the sex of the newborn baby (Table 1).

**Fig. 1:**
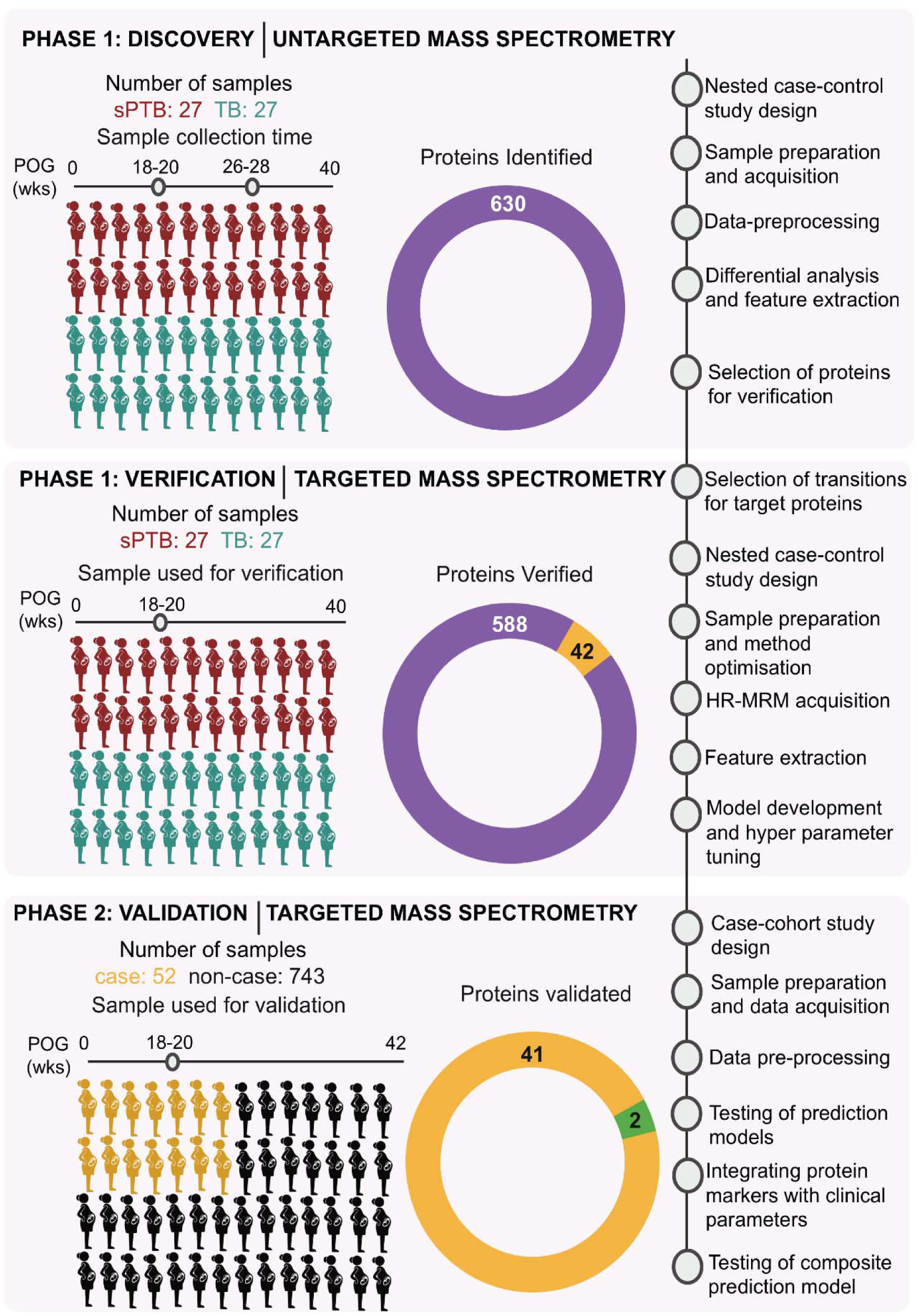
Overall workflow of the study. The study was divided into two Phases. Phase 1 consisted of discovery and verification arms, which included the untargeted and targeted proteomics of the plasma samples from 54 mothers, along with the development of the prediction models. Phase 2 involved large-scale validation of selected plasma proteins and prediction model testing in a case-cohort study design to evaluate the clinical importance of markers.

**Fig. 2:**
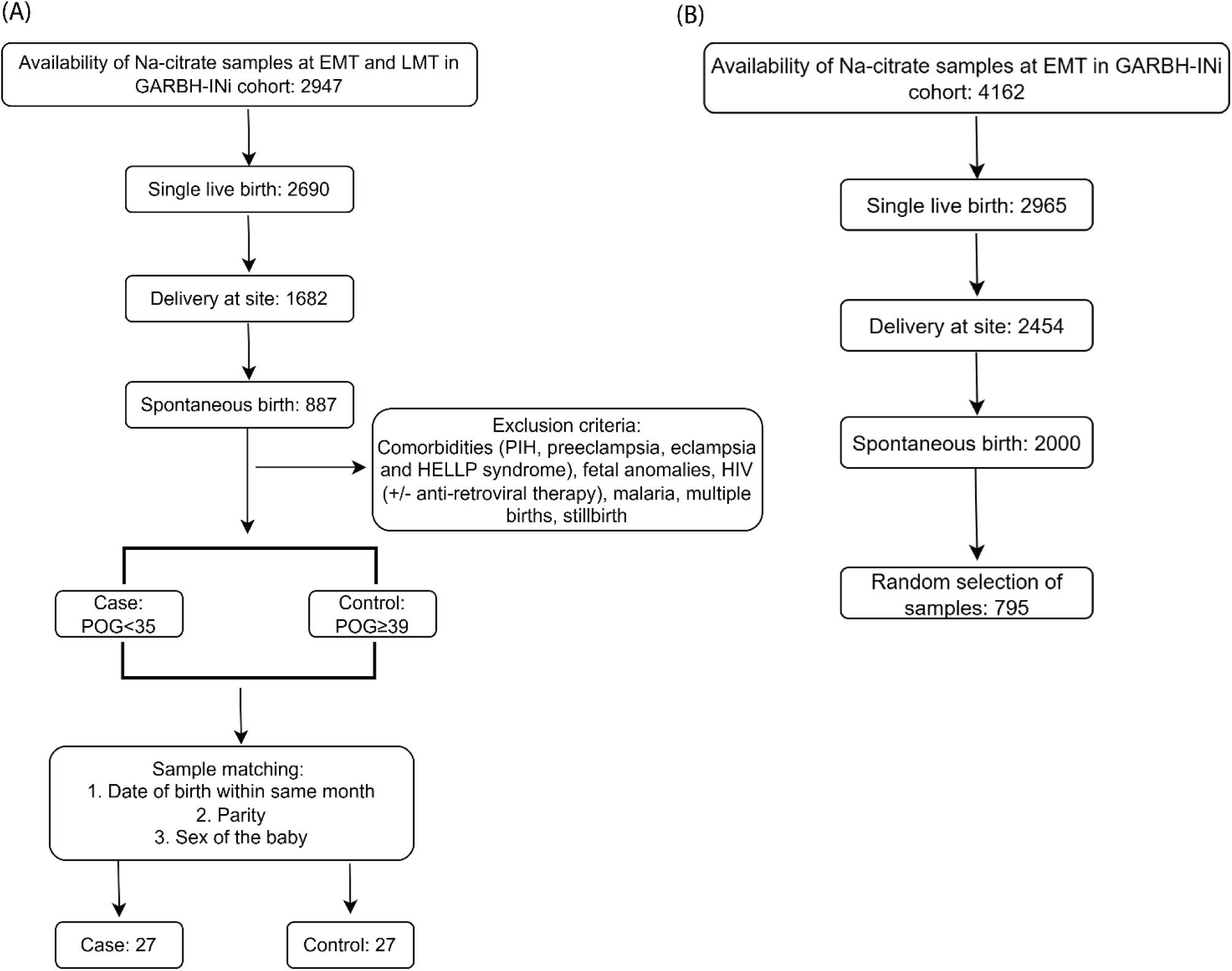
Sample selection workflow: (A, B) Flow chart depicting the sample selection criteria for Phases 1 and 2, respectively.

**Table 1:** Clinical and demographic characteristics of participants in Phases 1 and Phase 2 at baseline and delivery.

| Characteristic | Participants in Phase 1 (n=54) <sup>1</sup> | Participants in Phase 2 (n=795) <sup>2</sup> |
| --- | --- | --- |
| Age | 23 (21,25) | 23 (21,26) |
| Body mass index | 19.6 (17.6, 22.9) | 20.8 (18.4, 23.6) |
| Body weight (in Kgs) | 43 (40, 53) | 49 (43, 56) |
| POG at delivery | 36.9 (32.7, 39.4) | 39 (37.86, 39.86) |
| Participant education |  |  |
| College | 23 (43%) | 340 (43%) |
| School | 9 (17%) | 338 (42.8%) |
| Illiterate | 22 (41%) | 117 (15%) |
| Sex of the baby |  |  |
| Male | 28 (55%) | 370 (50%) |
| Female | 23 (45%) | 370 (50%) |
| Unknown | 3 | 55 |
| Parity |  |  |
| 0 | 26 (48%) | 421 (53%) |
| 1 | 21 (39%) | 269 (34%) |
| 2 | 5 (9.3%) | 85 (11%) |
| 3 | NA | 17 (2.1%) |
| 4 | 2 (3.7%) | 2 (0.3%) |
| 5 | NA | 1 (0.1%) |
| Participant occupation |  |  |
| Professional | NA | 3 (0.4%) |
| Skilled | NA | 21 (2.6%) |
| Unemployed | 53 (98%) | 736 (93%) |
| Unskilled | 1 (1.9%) | 35 (4.4%) |

<sup>1</sup>A total of 54 plasma samples were selected for Phases 1 study whereas <sup>2</sup>795 samples were selected for Phase 2. The table represents the sociodemographic and clinical variables of the selected participants, both at baseline and at time of delivery. Data are expressed as median (Q1, Q3) or number (percentage), counts as *n* (%) for continuous and categorical outcome variables, respectively.

In Phase 2, from a defined universe consisting of live, singleton, spontaneously delivering mothers with availability of plasma samples at 18-20 weeks of POG, 795 mothers were randomly selected in a case-cohort study design. Among the selected participants, those who delivered live births at less than 35 weeks of POG with spontaneous onset of labor or preterm premature rupture of membrane or cervical insufficiency were considered as cases, whereas the rest were kept in the non-case group and were used as comparators (Fig.1 and 2B). Median age and body weight of the mothers at first ANC visit were 23 years (IQR: 21, 26) and 49 kg (IQR: 43, 56), respectively. Similar to participants of phase 1, around 50% of the women in phase 2 were also nulliparous (Table 1).

### Temporal characterization of the global plasma proteome across mid-trimester (Phase 1-Discovery)

For discovery proteomics, neat plasma digested peptide samples were subjected to bottom-up proteomics in Zeno SWATH DIA mode on a ZenoTOF 7600 mass spectrometry system. We identified 4387 peptides corresponding to 635 protein groups (710 proteins) in EMT samples and 4379 peptides mapped to 632 protein groups (712 proteins) in LMT samples. Mass spectrometry data acquisition achieved an overall data completeness of around ∼ 85%, with an average of 4.2 data points per peak and ∼7.5 peptides per protein group in neat plasma samples. The median precursor percent coefficient of variation (%CV) in the sPTB and term birth (TB) groups of EMT samples was 35.12 % and 37.16 %, respectively (Fig. S1A). Whereas, in the LMT group, the median %CV was 34.95 % and 36.49 % in the sPTB and TB groups, respectively (Fig. S1B).

Global normalization, transformation, and imputation were performed during data pre-processing. The intensity distribution of non-imputed as well as imputed data was found to be Gaussian in both EMT (Fig. S1C) and LMT (Fig. S1D) samples, reflecting that the missing value imputation did not change the overall data distribution. Mean (IQR) and mean of the median intensity values for EMT samples were 3.27 (sd: 0.079) and 6.72 (sd: 0.18), respectively (Fig. S1E, S1G). Similarly, for LMT samples, the mean (IQR) was 3.27 (SD: 0.086), and the mean median intensity values were 6.72 (SD: 0.18) (Fig. S1F, S1H). Principal component analysis (PCA) separated the sPTB and TB samples into two distinct but partially overlapping islands, with a maximum variation of 19.5 % at PC1 and 7.9 % at PC2 in the EMT group (Fig. 3A). In LMT samples a similar pattern of sample clustering was observed in PCA with maximum variation of 16.7 % at PC1 and 9% at PC2 (Fig. 3B). High measurement precision and a robust biological distinction between sPTB and TB were supported by PCA-based sample clustering, along with comparatively low precursor %CV, a narrow interquartile range, and minimal standard deviation in sample medians.

**Fig. 3:**
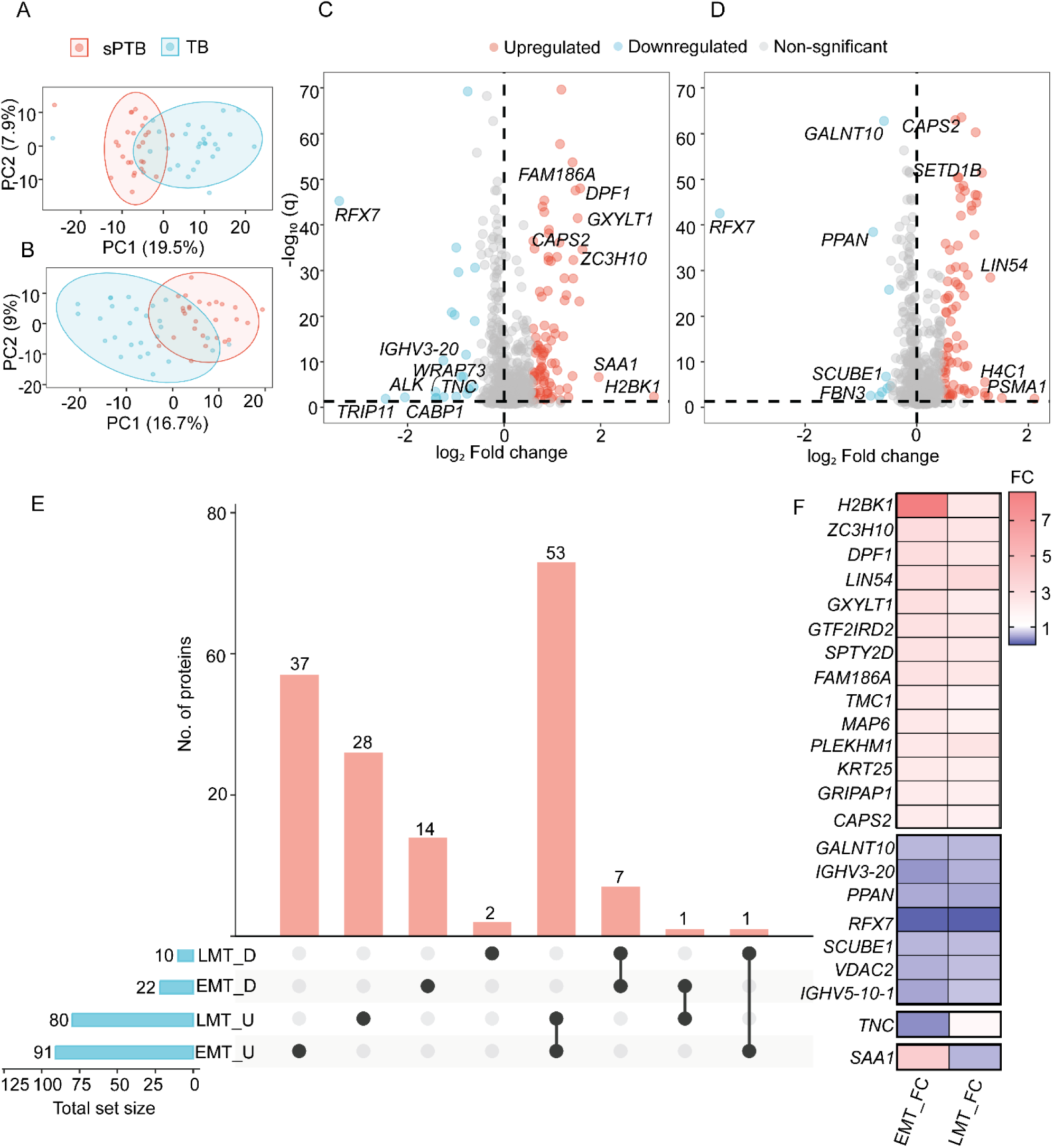
Differential expression of plasma proteome in sPTB outcome. **(A, B)** The proteomics data of 54 samples were plotted on the PCA plot using PC1 and PC2 components for EMT and LMT samples, respectively. The percentage variance for each principal component is mentioned in brackets. **(C, D)** DEPs between sPTB and TB groups at the EMT and LMT stages were plotted in the form of a volcano plot, respectively. The horizontal dotted line represents the -log_10_ (q) value cutoff of 1.3, whereas vertical lines mark the zero-fold change point separating the proteins regulated in opposite directions. Significant DEPs were highlighted with colors, and a few of them were annotated with the gene name. **(E)** The common and unique DEPs between the EMT and LMT samples were represented in the form of an Upset plot. **(F)** The fold change of selected DEPs common to sPTB and TB groups was plotted as a heatmap. Statistical evaluation of the DEPs was done using unpaired two-tailed students’ t-test with multiple testing correction.

### Plasma proteome alterations precede the onset of sPTB

Differential protein expression analysis (FDR-adjusted p<0.05, absolute log_2_ FC cut off: 0.58) between the sPTB and TB group displayed 113 and 90 significantly altered proteins in EMT and LMT plasma samples, respectively (Fig. 3C, 3D, Tables S1 and S2). Many DEPs were upregulated (EMT: 91, LMT: 80) and only a few proteins were found to be downregulated in sPTB plasma collected at both points (EMT: 22, LMT: 10). Among the upregulated proteins, 53 were common to both EMT and LMT, 37 were unique to EMT and 28 were unique to LMT. Similarly, among the down-regulated proteins, 7 were common to both and 14 were unique to EMT, and 2 were unique to LMT (Fig. 3E). Notably, approximately 97% of the common modulated proteins displayed consistent regulatory trends in both EMT and LMT, except for *TNC* and *SAA1* proteins, as illustrated in the heatmap (Fig. 3F).

The protein-protein interaction (PPI) map of the DEPs obtained from the String interaction database divided the differential proteins of the EMT group into 17 subnetworks. Of these, major sub-network 1 consisted of 58 seed proteins connected to 1300 nodes via 1550 interactions, as depicted by the total number of edges. The second biggest network obtained was sub-network 2, which mapped 2 seed proteins interacting with 59 nodes via 68 edges. Similarly, for the DEPs identified in the LMT group, 19 sub-networks were extracted from the PPI map. Out of 90 DEPs, major sub-network 1 mapped 41 proteins connected to 1340 nodes, revealing 1587 interactions (Table S3). The interplay between the DEPs and their possible interactors in the major sub-networks was mapped into various functional modules using the InfoMap algorithm integrated with NetworkAnalyst. Functional enrichment of modules in the EMT set revealed their association with notable proteins including NF-kB1, Fibroblast growth factor receptor 4 (FGR4), Platelet derived growth factor beta (PDGFRB), Inositol 1,4,5-Trisphosphate Receptor Type 1 (ITPR1), Microtubule Associated Protein 6 (MAP6), Inner Centromere Protein (INCENP) which enrich for molecular functions such as cytokine receptor binding, growth factor and hormone receptor activity, calcium ion binding, and histone acetyltransferase activity (Fig. 4A). The functional modules of the major sub-network mapped with DEPs from the LMT set enriched molecular functions, including calcium ion transmembrane transporter activity, ubiquitin binding, oxidoreductase activity, growth factor activity and GTPase activity (Fig. 4B).

**Fig. 4:**
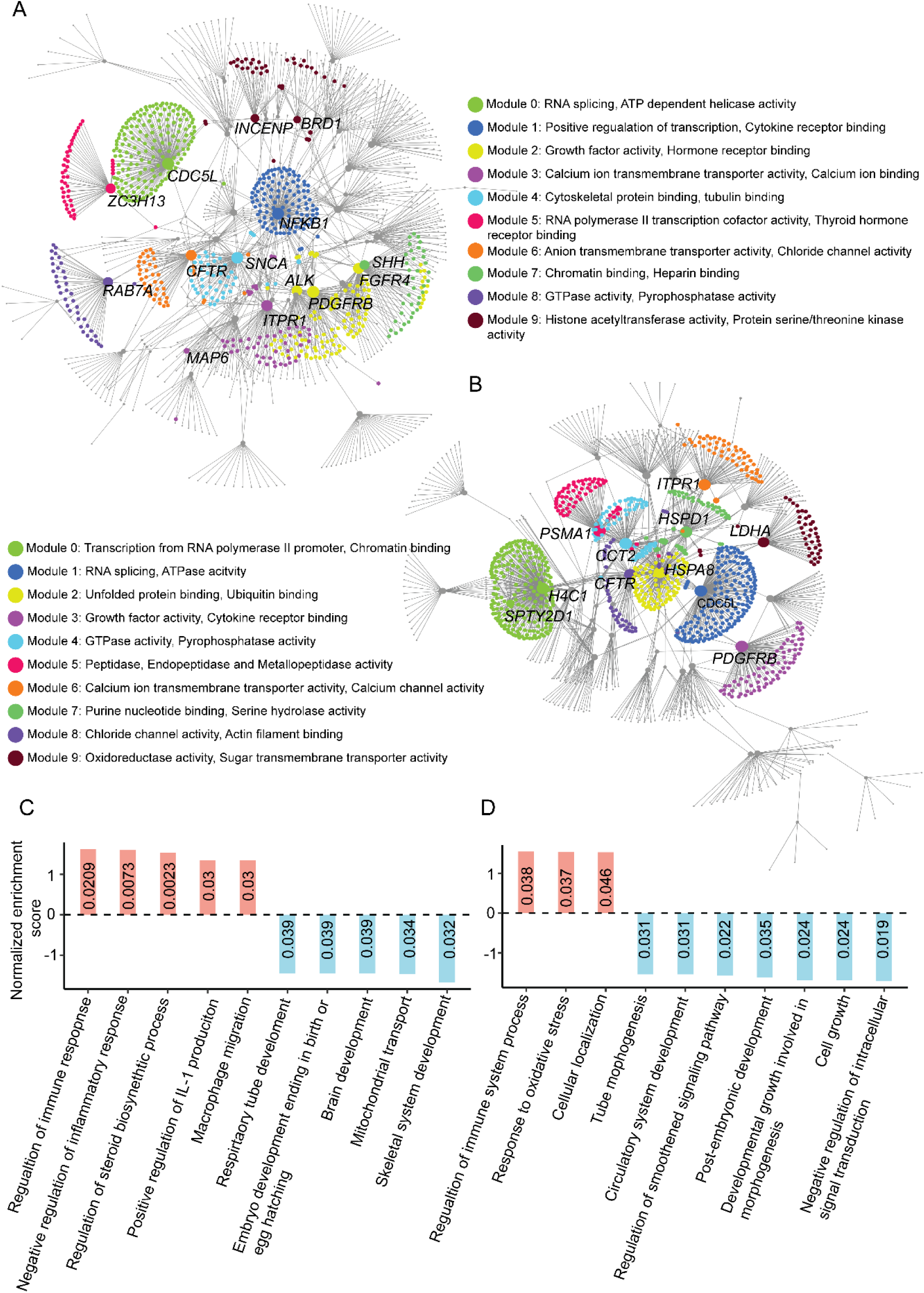
Pathway enrichment of DEPs. **(A, B)** Sub-network corresponding to the PPI interaction map of DEPs in EMT and LMT samples. Top modules characterized based on their functional interaction were represented with different colors, and the molecular functions associated with each module were mentioned. Seed node of each module was annotated with the gene name. **(C, D)** GO: BP pathway enrichment of DEPs at the EMT and LMT stages. Pathways with positive normalized enrichment scores show upward regulation in the sPTB group, whereas those with negative scores show downregulation. Corresponding p-values are mentioned in each bar.

The Gene Set Enrichment Analysis (GSEA) using the GO: BP database demonstrated that development-associated biological pathways, including skeletal system development, brain development, embryo development, and respiratory tube development, were negatively regulated in the sPTB samples as compared to term samples collected at EMT. These important pathways were associated with downregulation of specific proteins such as Sonic hedgehog (*SHH*), Bone morphogenetic protein (*BMP8B*), Thyroid receptor interacting protein 11 (*TRIP11*), Alk tyrosine kinase receptor (*ALK*), Telomeric repeat binding protein (*TERF2*) and Tenascin (*TNC*). In contrast, pathways such as steroid biosynthesis, macrophage migration and positive regulation of IL-1 production were upregulated in this group (Fig. 4C). A similar downregulation in development-related terms, such as post-embryonic development, tube morphogenesis, developmental growth involved in morphogenesis, and circulatory system development, was observed in sPTB samples collected at LMT timepoint. Regulation of the immune system, response to oxidative stress, and cellular localization were upregulated pathways in this sample set (Fig. 4D).

### Differentially expressed proteins demonstrate predictive power for the early detection of sPTB

To identify early-stage potential biomarkers for sPTB and develop a prediction model, we employed two different feature extraction algorithms, namely RFE-CV and SHAP, fitted with three different classifiers viz. support vector machine (SVM), logistic regression (LR), and random forest (RF) to filter through the large number of proteins quantified in the plasma samples collected at the early mid-trimester stage of pregnancy in the discovery Phase. We leveraged the power of RFE with the five-fold cross-validation to eliminate the less significant features and select a protein subset with enhanced predictive efficiency. When fitted with the SVM classifier, a set of 8 features (mean_test_score: 0.945) was selected, whereas a set of 12 (mean_test_score: 0.963) and 73 (mean_test_score: 0.945) features were selected with LR and RF classifiers, respectively (Fig. 5A). With SHAP algorithm, we were able to intuitively understand the impact of individual proteins on the likelihood of spontaneous preterm delivery because of its differential expression in the plasma samples. RFX7 had the highest effect on model output in both SVM and LR classifiers, and the participants with lower expression of RFX7 at the early mid-trimester stage were more likely to have sPTB. RUNDC3A had the maximum predictive power for sPTB with the RF model. We chose the top 20 proteins from all three classifiers which have the highest influence on model output, as listed in the SHAP plot (Fig. 5B). To strengthen our protein selection strategy, we overlapped the ML-extracted proteins with DEPs and identified 42 common proteins (Fig.6A). Among these, 5 proteins, namely IGHV3-20, IGLV2-8, PPAN, RFX7, and TERF2, were downregulated in the sPTB plasma, while the rest were upregulated as depicted by discovery proteomics data (Fig. 6B).

**Fig. 5:**
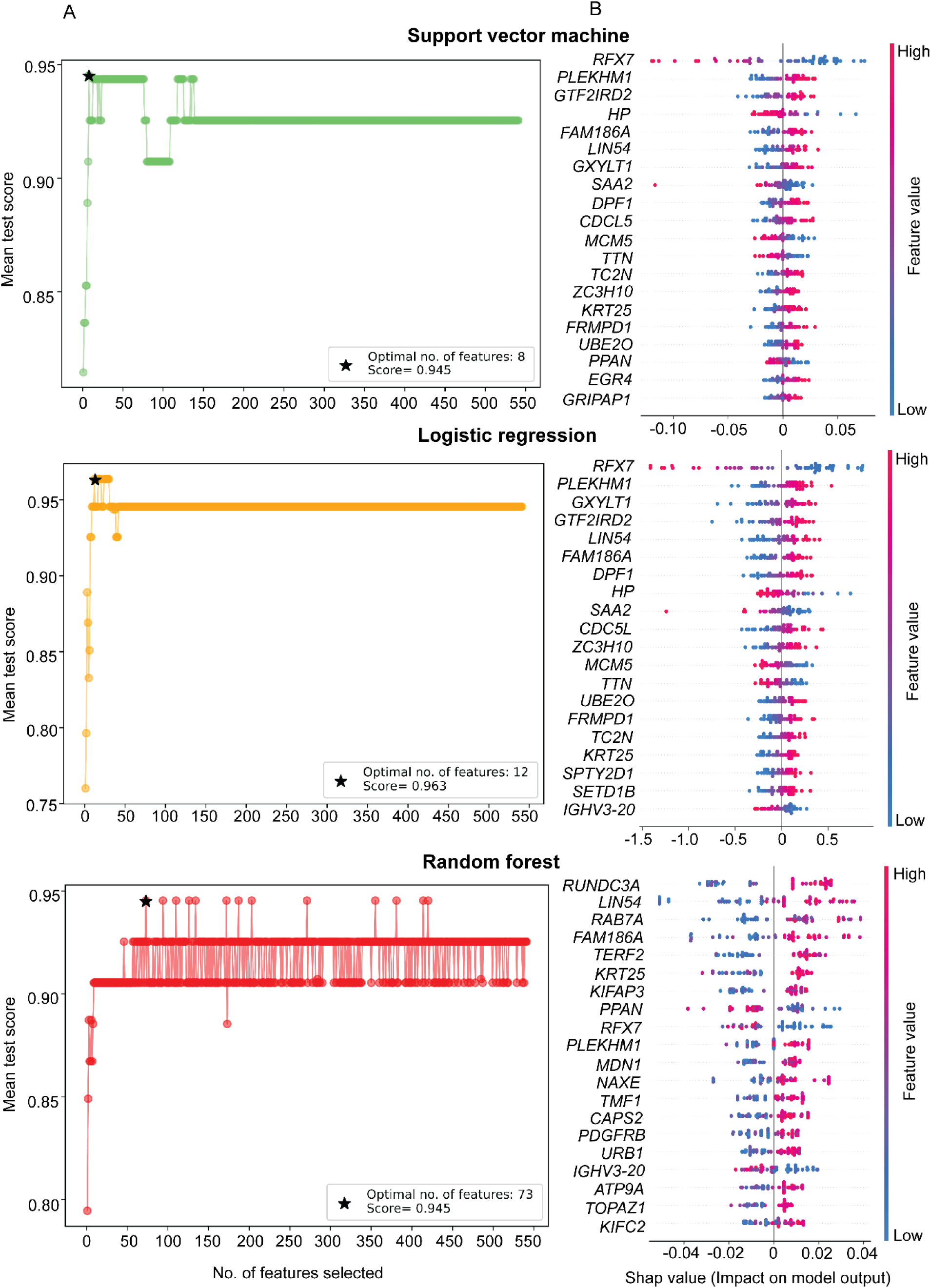
Selection of predictive proteins for protein filtration. **(A)** A dot plot representing the mean test score of each subset generated by the RFE-CV algorithm fitted with different classifiers. Star marks that sub-set which consists of proteins selected by the RFE-CV with highest score; its accuracy score is mentioned in the legend. **(B)** SHAP visualization plot of the top 20 proteins in terms of their impact on the model output. The spread of the dots is proportional to the contribution of protein in predicting sPTB. Color represents the level of proteins in the plasma, which is denoted by a gradient from blue (low) to red (high). The positioning of the dots with respect to the middle line indicates the chances of a sPTB (right side) or TB (left side) outcome due to proteins’ expression in plasma.

**Fig. 6:**
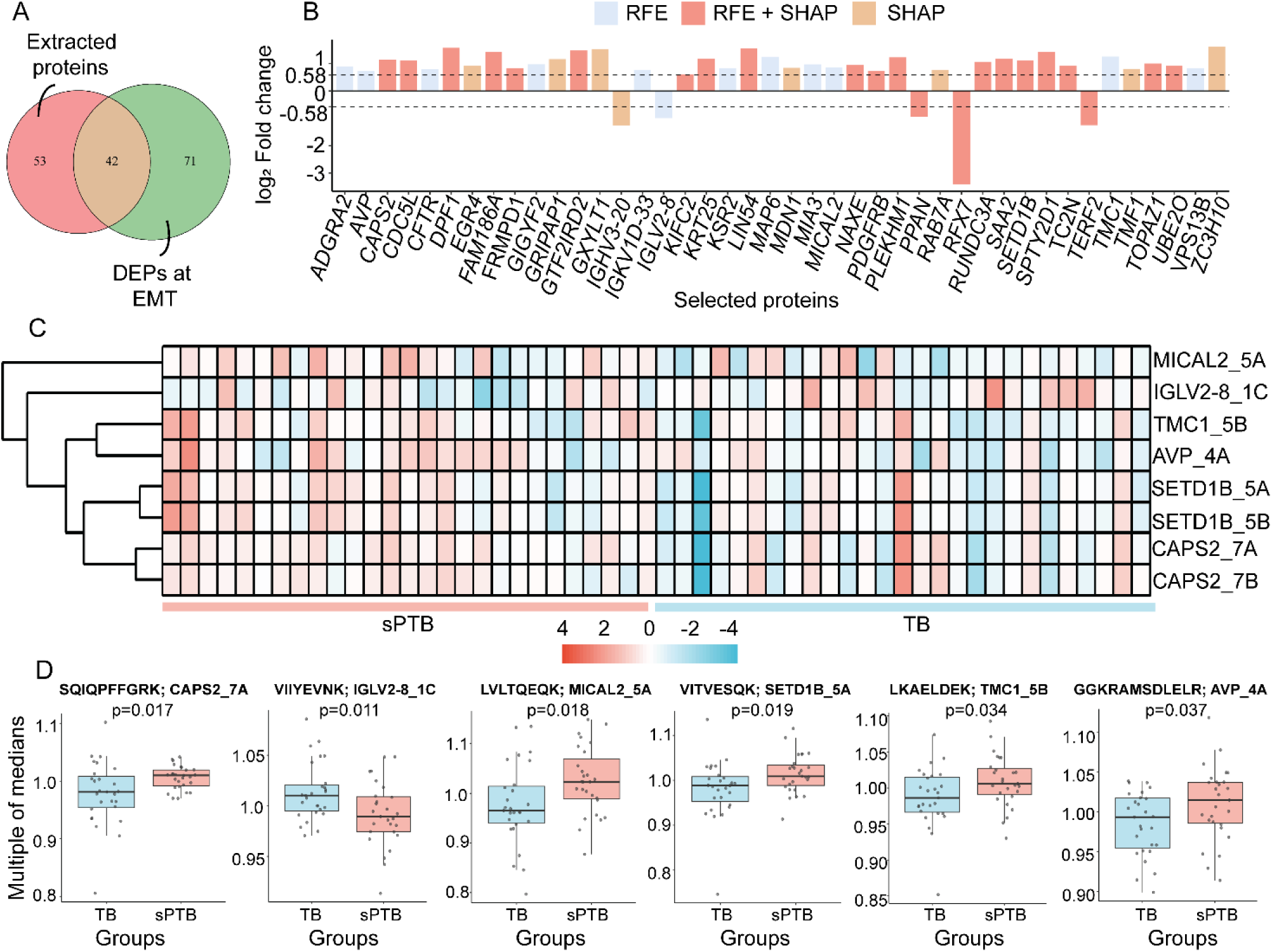
Targeted measurement of proteins for verification. **(A)** 42 proteins were found to be common between the DEPs and ML extracted proteins. **(B)** All the selected proteins were plotted in a bar plot representing their log_2_ Fold change. The color of the bar denotes the algorithm used to extract a particular protein. **(C)** Multiple of medians (MOM) of statistically significant transitions with direction of regulation the same as that of discovery data were presented in a heat map. **(D)** Box and whiskers plots displaying at least one statistically significant transition for each protein. The overlaid scatter plot denotes individual measurements. The whiskers represent minimum and maximum, whereas boxes show the 25^th^ percentile, median, and 75^th^ percentile. Regulation between two groups was compared using an unpaired two-sided Wilcoxon rank sum test. p-values for the individual tests are mentioned. Transitions corresponding to a particular peptide of a protein are annotated above the plot along with the peptide sequence. The number after underscore in the annotation is the peptide number and alphabet represent the chosen transition.

### Targeted measurement of proteins selected from the discovery phase plasma proteomics analysis (Phase 1-Verification)

Next, we designed an MRM^HR^ workflow for 42 proteins selected from the discovery proteomics to verify their differential expression pattern using a targeted pipeline. These measurements were carried out in the phase 1, 27 pairs of nested case-control sample sets. Extracted peaks were manually curated, and peak areas of each product ion/ transition were exported. Raw intensities were normalized using the median-centric method from the proBatch package (Fig. S2A, S2B). Statistical evaluation of individual transitions was done using an unpaired two-sided Wilcoxon rank sum test, identifying 14 transitions corresponding to 10 proteins as significantly regulated between the sPTB and TB groups. Out of these 14 transitions, the direction of regulation of 8 transitions was found to be the same to their corresponding proteins’ regulation in the discovery proteomics differential analysis; these transitions corresponded to the following proteins-SET Domain Containing 1B, Histone Lysine Methyltransferase (*SETD1B*), MICAL2, Arginine Vasopressin (*AVP*), Transmembrane Channel Like 1 (*TMC1*), CAPS2 and Immunoglobulin Lambda Variable 2-8 (*IGLV2-8*). Except for IGLV2-8, transitions of all five other proteins were upregulated in the sPTB condition (Fig. 6C, 6D). To further strengthen our protein selection pipeline at the targeted level as well, we used the Boruta feature extraction method to extract the transitions predictive of sPTB. Five transitions were extracted with z-score importance higher than the shadowMax, out of which Boruta confirmed CAPS2_7A, CAPS2_7B, MICAL2_5A, and TMF1_3C, whereas GIGYF2_3D was selected as a tentative feature (Fig. 7A). Of the four confirmed transitions CAPS2_7A, CAPS2_7B, MICAL2_5A were also statistically significant on unpaired two-sided Wilcoxon rank sum test and were further selected for large scale validation as a part of phase 2 (Fig. 1).

**Fig. 7:**
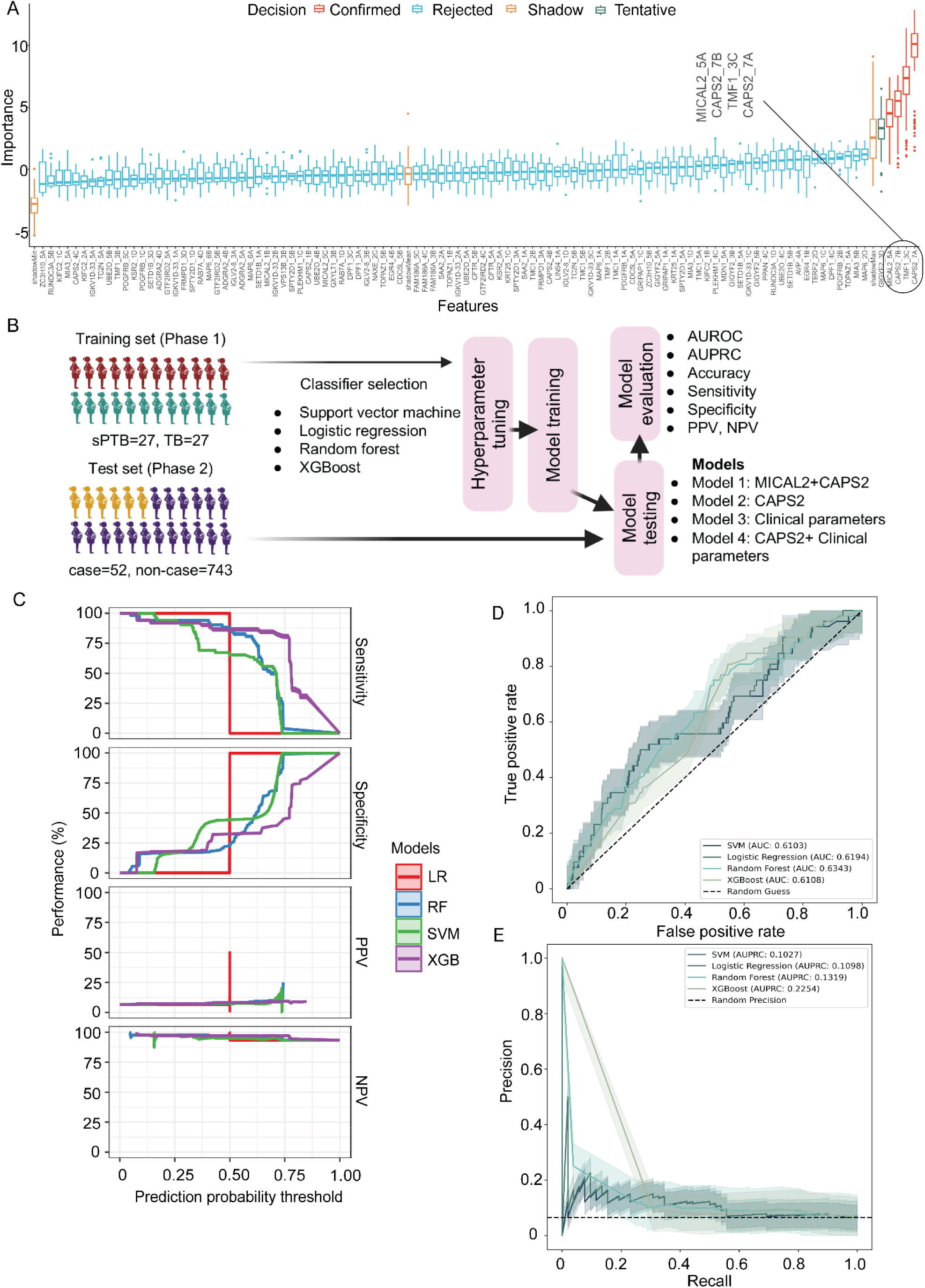
Targeted validation and development of prediction model. **(A)** A Boruta visualization plot displaying the z-score importance of confirmed, tentative, rejected, and shadow features. The inset shows transitions confirmed by Boruta. **(B)** Schematic of prediction model development and testing pipeline. **(C)**Prediction performance of Model 2 developed using different classifiers depicted by sensitivity, specificity, PPV and NPV calculated at multiple prediction probability cutoffs on the test set. **(D, E)** Receivers operating curve and precision recall curve show the performance of individual models built using plasma *CAPS2* levels only. AUROC and AUPRC are mentioned in the lower right and upper right of the plots respectively. Shaded regions represent the 95% confidence interval calculated by performing 1000 iterations. PPV: Positive predictive value, NPV: Negative predictive value

### Targeted validation identified *CAPS2* as a potential predictor of spontaneous preterm birth cases

In Phase 2, we validated the signatures from the verification phase using a targeted mass spectrometry pipeline. A randomly selected sub-cohort of cases (POG<35) and the rest of the non-case participants were used for targeted validation (Fig. 2B). An MRM^HR^ workflow was designed to measure the expression of CASPS2 and MICAL2 proteins in 795 plasma samples. The raw data were acquired, manually curated, and further normalized using the median-centric method (Fig. S2C, S2D). To evaluate the power of these candidate protein markers for predicting a future case, we developed the models using the pipeline presented in Figure 7B. Models were trained on verification samples and tested on the validation set. Model 1, based on CAPS2 and MICAL2 expression, achieved an AUC ranging from 0.52 to 0.56 on the ROC curve (Fig. S3B) and 0.07 to 0.0747 on the PRC curve (Fig. S3C) using four different classifiers. As depicted in Figure S3A, we evaluated the prediction performance of the model across a range of prediction probability-cutoffs.

Across all four classifiers, the sensitivity decreased at higher cutoffs, whereas the specificity increased, representing the expected trade-off. Positive predictive value (PPV) settled between 6%-8% across the threshold; except with LR and XGB classifiers, the negative predictive value stayed more the 90% (Table S5).

In a previous study on shared genomic sequence analysis of SNP genotyping, Workalemahu *et al.* reported that the sPTB-delivering women shared a chromosome locus 12q21.1-q21.2 in two independent pedigrees. They discussed that this chromosomal region corresponded to genes CAPS2 and KCNC2 ^18^. This report motivated us to build a prediction model using the expression of CAPS2 transitions only (Model 2). The risk of incident sPTB was predicted with an improved AUROC of ranging from 0.61-0.63 (Fig. 7D) and an AUPRC of 0.10-0.22 on different classifiers as compared to Model 1(Fig. 7E). The trends of model performance are visualized in Figure 7C, which displays how each metric changes with the decision threshold, aiding in selecting an appropriate cutoff for practical use. At a prediction threshold of 0.4 with the XGBoost approach of Model 2, the sensitivity to predict sPTB was high at 90.4% (95% CI: 81%-98%) with a specificity of 25.2% (95% CI: 22%-28%). The other approaches of the models, including support vector machines, logistic regression, and random forest, had similar and marginally lower sensitivity and specificity profiles (Table S6).

Next, we attempted to develop a comprehensive prediction model by combining the protein biomarker candidate *CAPS2* with the clinical parameters of the mothers. Metadata from the Garbh-Ini cohort was used. We re-employed the RFE-CV method to select a subset from the clinical parameters with the highest predictive power for sPTB. We achieved a maximum accuracy score of around 0.6 from a subset consisting of 12 parameters (Fig. S4A). We further sorted them based on their importance for prediction tasks; the use of contraceptives before current pregnancy was ranked highest in predicting sPTB, followed by the use of unsafe toilets and exposure to tobacco. Previous reports have discussed that hemodynamic changes associated with uterine arteries in the early stages of pregnancy are associated with adverse pregnancy outcomes^19^. Our data also highlighted that right and left uterine artery pulsatility index measurements can be employed as a predictive marker for sPTB (Fig. S4B). Employing 4 different classifiers, we achieved a maximum AUC of 0.58 (95% CI: 0.52-0.65) on the ROC curve (Fig. S4D) and 0.0778 (95% CI: 0.055-0.11) on the PRC curve with SVM classifier (Fig. S4E) when the model was trained and tested with selected clinical and demographic parameters (Model 3). Combining these parameters with CAPS2 plasma expression (Model 4) didn’t improve the predictability beyond Model 2, and it reported the best AUROC of 0.59 only (Fig. S5B). The prediction performance trends of Model 3 and Model 4 across decision thresholds are depicted in Figure S4C and S5A and Table S7 and S8, respectively.

### CAPS2 exhibits enhanced predictive ability for identifying high-risk sPTBs

PTB can be divided into different categories based on the week of delivery. With each additional week of gestation, the risk of neonatal mortality and major and minor morbidities decreases. In a cohort-based study, it has been reported that beyond 32 weeks of gestation, the chances of survival of neonates without any co-morbidities become more than 30%, along with a significant reduction in hospital stays of babies by 8-11 days^20^. Thus, to leverage the predictive power of *CAPS2* expression-based model 2 for high-risk sPTB cases, we re-categorized our validation cohort into cases (POG ≤ 32 weeks, n=31) and the rest as controls (n=764, Fig. 8A). The performance of Model 2, when tested on high-risk sPTB cases, improved as indicated by increased AUROC. Its value ranges between 0.63-0.71 for the different classifiers employed (Fig. 8C), although the AUPRC on the PRC curve (Fig. 8D) remained mostly similar. The prediction performance was better for high-risk sPTB, with the XGBoost approach of Model 2 achieving a sensitivity of 90.3% (95%CI: 78%-100%) for a specificity of 39% (95%CI: 36%-42 %) at a prediction threshold of 0.7 (Fig. 8B and Table S9).

**Fig. 8:**
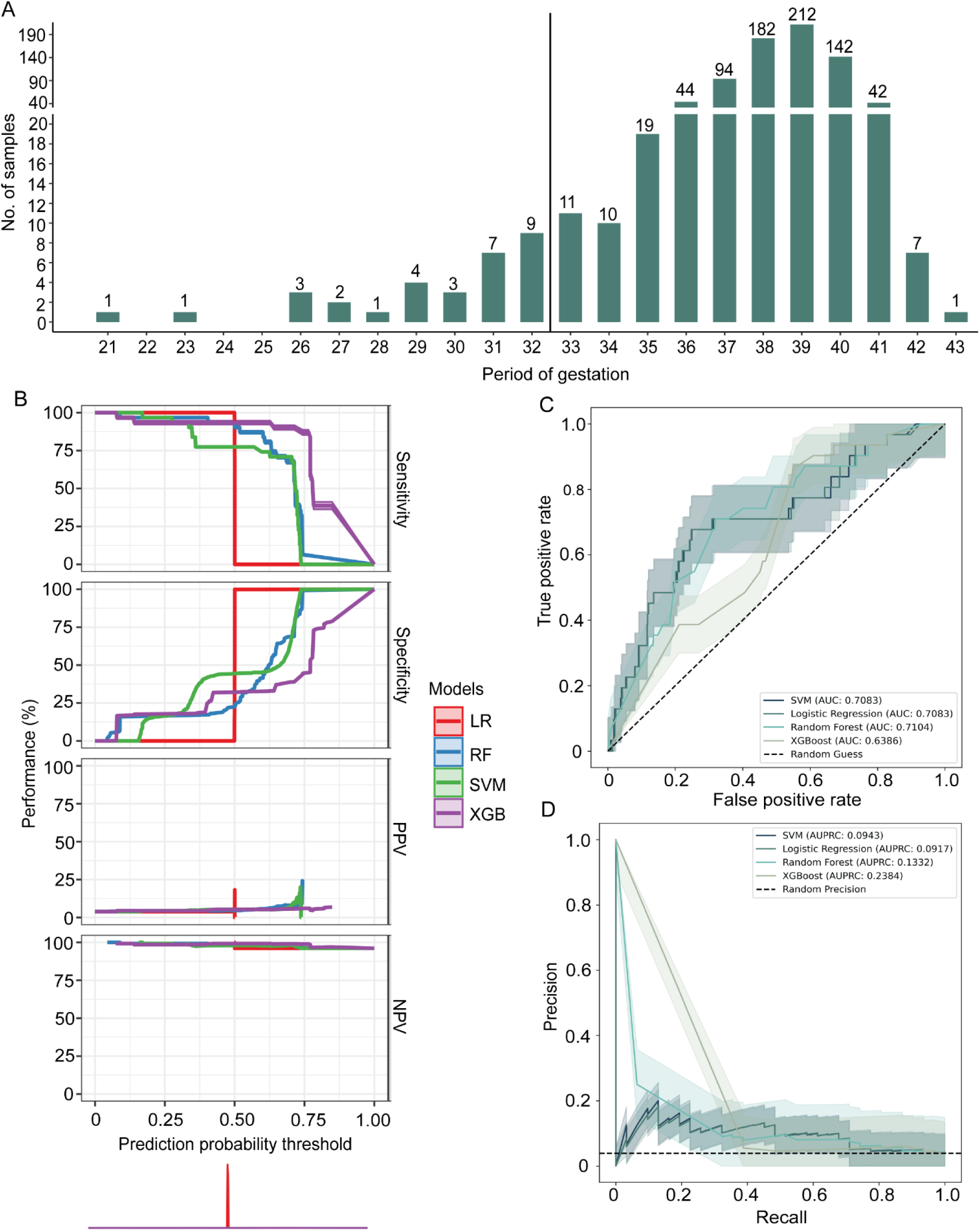
Predictive capability of *CAPS2* in high-risk sPTB cases. **(A)** Number of samples available in each POG group in the validation sample set. **(B)** Prediction performance of Model 2 when tested in high-risk sPTB cases depicted by sensitivity, specificity, PPV and NPV calculated at multiple prediction probability cutoffs **(C, D)** Receivers operating curve and precision recall curve shows the improved predictive performance of the model for sPTB cases delivered at ≤32 weeks of POG as depicted by AUROC and AUPRC mentioned in the lower right and upper right of the plots respectively. Shaded regions represent the 95% confidence interval calculated by performing 1000 iterations. PPV: Positive predictive value, NPV: Negative predictive value

## Discussion

Preterm birth complications are the leading cause of neonatal death worldwide. The sPTB, which includes both idiopathic PTB and preterm premature rupture of membranes (PPROM), accounts for 70-75 % of the total PTB tally^21^. Despite significant advances in biomedical research, major gaps persist in our understanding of the molecular mechanisms underlying spontaneous preterm birth, and no sensitive or reliable biomarker is yet available for the timely identification of mothers at risk. We undertook a global proteomics assay on a nested case-control sample set of sPTB and identified approximately 630 protein groups with dependable data completeness from the neat plasma, which were benchmarked with the previously published plasma/serum proteomics reports^22^. We were able to map the candidate biomarkers for sPTB reported in the literature, such as Fetal fibronectin, C-reactive protein, β-human chorionic gonadotropin, Pappalysin-1, Pregnancy-specific β-1-glycoprotein 1, Desmoplakin^23^ and Serum Amyloid A^9^, in both EMT and LMT samples, till the protein group rank of more than 600, highlighting the reliable depth of our proteome dataset in the context of the pathology. We found that ∼16% of the measured plasma proteome’s expression differed from its baseline values at both early and late mid-trimester stages, resulting in a deviation from the expected trajectories of changes in protein levels during normal pregnancy^24^. The second trimester is an important stage for the development and maturation of the fetuses’ organs^25^. GO: BP enrichment established that the regulation of critical pathways related to the development of the respiratory tube, brain, circulatory system, and skeletal system was disturbed due to changes in the plasma proteome. Additionally, crucial molecular functions, including calcium transmembrane transport, ubiquitin-binding, growth factor and oxidoreductase activity, were impaired.

Employing machine learning-backed feature extraction methods, we identified 42 proteins with considerable predictive power. We designed a targeted assay and finally narrowed it down to two proteins-*MICAL2* and *CAPS2* for independent validation in a larger sample set in a case-cohort study design. *MICAL2* is a monooxygenase that promotes the depolymerization of F-actin and manages the cellular remodeling associated with it^26^. This protein has been extensively studied with respect to cancers, such as pancreatic ductal adenocarcinoma (PDAC), where it is reported to promote the activity of ERK1/2 and AKT, affecting cell growth, proliferation, and differentiation^27^. Ismail et al. demonstrated that an imbalance in ERK and AKT activity is a major factor in unexplained first-trimester recurrent miscarriages in Egyptian women^28^. We argue that the upregulated *MICAL2* in the mid-trimester plasma observed in our study is an indicator of aberrant ERK1/2 and AKT protein activity that can contribute to adverse pregnancy outcomes such as PTB. SNP genotyping carried out on blood samples from women who experienced sPTB identified a chromosomal region annotated to the *CAPS2* gene as a conserved genetic element shared among the mothers within two independent pedigrees^18^. Calcyphosin 2 is a Ca^2+^ binding protein consisting of 4 EF-hand motifs and is phosphorylated by cAMP-dependent kinase^29^. Calcium is a critical macronutrient for the maintenance of a healthy pregnancy, having important roles in muscle contraction, bone formation, enzyme and hormone function^30^. A decrease in the serum calcium concentration is reported as a potential predictor of preterm deliveries at 32 weeks of gestation^16^. Low levels of calcium in the blood have the potential to stimulate the release of parathyroid hormone, leading to increased intracellular calcium levels, followed by smooth muscle contraction, resulting in premature labour^30^. Thus, changes in the levels of plasma calcium-binding proteins can induce spontaneous preterm labor by regulating uterine muscle contractions.

The prediction model developed herein using the XGBoost classifier was powerful enough to predict the sPTB cases with a sensitivity above 90%, especially the high-risk sPTB cases (POG≤ 32 weeks), with a reasonable specificity. We expect Model 2, based on *CAPS2* expression, to be a good candidate as a screening biomarker for the prediction of high-risk sPTB. We are planning to evaluate this marker in combination with other clinical, ultrasound, and multi-omics-based predictors, both cross-sectionally in the mid-trimester and longitudinally along the three trimesters of pregnancy.

From a public health perspective, there are no currently existing screening tools for identifying women at high risk of sPTB at the primary and secondary care levels. This single marker screening test using the Calcyphosine 2 protein will be able to identify 90% of women at risk of spontaneous preterm birth while avoiding unnecessary expensive tests in 40% of low-risk women. When coupled with other prediction tools such as ultrasound and metabolomics tools, we anticipate *CAPS2* to enhance the overall prediction of sPTB.

In this study, we leveraged the power of large-scale proteomics to quantify the global plasma proteome of sPTB-delivering women for the first time. Our observations established that levels of specific proteins in plasma fluctuate steadily through the mid-trimester as a pivotal modulation that could be causative of future spontaneous delivery. Targeted measurements and ML extractions identified *CAPS2* as a candidate marker that could predict the sPTB with high sensitivity. The outcome of our prediction model is strengthened by our validation study design. We identified aberrant expression of *CAPS2* in sPTB samples matched to controls during the discovery Phase and subsequently used this set to train our predictive model. We successfully validated our model, accurately predicting sPTB in an independent case-cohort set, which not only included idiopathic deliveries but also samples from mothers with PPROM and associated comorbidities, such as pre-eclampsia. Such validation enhances the clinical relevance of the output and ensures its applicability in real-world settings. Our study has several important limitations. It represents development and internal validation effort; however, external validation is still required and is currently being considered across multiple regions of India. The available plasma sample metadata lacked certain clinical parameters, such as plasma calcium levels in women, which are physiologically relevant to *CAPS2* function; these could have been integrated to improve the model’s predictive performance. Additionally, investigating the underlying molecular mechanisms of the identified molecules was beyond the scope of this study.

In summary, we quantified approximately 635 protein groups in the neat plasma of pregnant mothers and identified prominent proteomic differences in those who delivered preterm at two distinct stages of the mid-trimester. These alterations were linked to dysregulated activity of several development-associated pathways, as well as processes involving calcium transport and GTPase activity. Notably, we identified *CAPS2* as a potential predictive biomarker for sPTB; when combined with additional biomarkers and validated in external cohorts, *CAPS2* holds promise for stratifying pregnant women into different risk categories for preterm birth. In the future, we aim to validate our findings in independent cohorts beyond the GARBH-Ini cohort to establish external validation. We also plan to integrate proteomics insights with metabolomics and transcriptomics profiles to generate comprehensive molecular profiling of maternal plasma and develop a more accurate prediction model for sPTB. Furthermore, we will investigate the molecular mechanisms underlying CAPS2 abnormal expression and its functional relevance in pregnancy outcomes.

## Materials and Methods

### Reagents

All general chemicals and reagents of LC−MS grade were purchased from Sigma-Aldrich (St. Louis, MO) and Fujifilm Wako chemicals (Richmond, USA).

### Study population and clinical definitions

The interdisciplinary Group for Advanced Research on Birth outcomes-DBT India Initiative (GARBH-Ini) has been conducting a prospective hospital-based pregnancy cohort in the Civil Hospital, Gurugram, Haryana, North India. This study was initiated with the hypothesis that time-series data collected on multidimensional characteristics, including clinical, imaging, environmental, genomic, epigenomic, meta-genomic, proteomic, and metabolomic parameters collected during pregnancy, can help identify women at high risk of delivering preterm (<37 completed weeks of gestation)^31^. The enrolled participants represent a mix of semiurban and rural populations attending the antenatal services of a secondary care hospital. Women with gestational age <20 weeks of gestation are enrolled and followed up to five times until delivery and once within 6 months postpartum, with visits aligned to the routine antenatal care schedule. As per study protocols, various biospecimens are collected, and ultrasound scans are performed at regular intervals.

Institutional Ethics Committees of all collaborating institutions approved this study. For phase 1 of the study, which consisted of discovery and verification arms, we used a nested case-control study design. The universe was defined, and 2947 eligible participants were selected from the cohort based on the following selection criteria: live, singleton, spontaneous birth with availability of plasma samples at EMT and LMT timepoints. Participants with comorbidities such as pregnancy-induced hypertension, preeclampsia, eclampsia, and HELLP syndrome, fetal anomalies, HIV (+/-anti-retroviral therapy), malaria, multiple births, and stillbirth were excluded from Phase 1 of the study. Of the remaining 887 participants, those who delivered spontaneously before 35 weeks of POG were classified as cases (spontaneous pre-term birth (sPTB)), while those who delivered at or after 39 weeks of POG were designated as controls (term birth (TB). For this study, controls were matched to baseline characteristics, except for the outcome. Matching criteria included delivery date (within the same month), parity, and sex of the newborn. A total of 27 participants were selected for each group (Fig. 2A).

For the validation Phase (Phase 2), the universe was defined by the following criteria: live, singleton, spontaneous birth, with available plasma samples at 18-20 weeks of POG. From this pool, 795 participants were randomly selected to form the sub-cohort. Within the sub-cohort, women who delivered live births before 35 weeks of POG due to spontaneous onset of labor, preterm premature rupture of membrane or cervical insufficiency were classified as cases. The remaining participants were categorized as non-cases and served as comparators (Fig. 2B).

### Plasma collection

Maternal venous blood samples were collected from pregnant women at 18-20 weeks and 26-28 weeks of gestation in sodium citrate-coated collection tubes. The samples were centrifuged at 2000 rpm for 20 min to isolate plasma. Separated plasma samples were mixed with protease phosphatase inhibitor (Halt protease phosphatase inhibitor, Thermo Scientific) and stored at -80°C until analysis.

### Sample preparation and protein digestion

The stored plasma samples were diluted twenty-fold in 100 mM ammonium bicarbonate (ABC), and 20 μL of the diluted neat plasma was aliquoted from each sample for further processing. Diluted plasma was further reduced with 10 mM dithiothreitol (DTT) at 56° C for 30 min and alkylated with 20 mM iodoacetamide (IAA), at room temperature (RT) for 1 h. The proteins were digested with mass spectrometry grade trypsin (1:15 *w/w*, Pierce, Thermo Fisher Scientific, Waltham, MA, USA), at 55 °C for 6 h. The digested peptide samples were next desalted using Oasis® HLB 1cc (10mg) extraction cartridges (Waters, Milford, MA, USA). The peptides were eluted using 200μl of elution reagent (70% acetonitrile and 30% water (v/v) with 0.1% formic acid). Eluted peptides were vacuum-dried and stored at -80°C until further use.

### Data-independent acquisition and processing (Phase 1-Discovery)

The vacuum-dried desalted plasma samples were resuspended in solvent A (98% H_2_O and 2% acetonitrile (v/v) with 0.1% formic acid) and injected into the ZenoTOF 7600 mass spectrometer (Sciex, MA, USA) coupled with ACQUITY UPLC M-Class system (Waters, MA, USA). 5µl digest from each sample was loaded, in triplicates, onto a Luna Micro Trap (20 x 0.3 mm, 5 µm, 100 Å, Phenomenex. Torrance, CA, USA) column at a flow rate of 5 μL/min for 3 min. The tryptic peptides were then eluted from nanoEasse^TM^ M/Z HSS T3 (100 Å, 1.8 µm, 300 µm x 150 mm, Waters, MA, USA) analytical column at a flow rate of 5 μL/min in a linear gradient of 5% solvent B (100% ACN (v/v) with 0.1% formic acid) to 80% solvent B in 27 min with a total run time of 32 min. A SWATH-DIA acquisition scheme of 65 overlapping variable windows covering a mass range of 350-1000 Da, with an accumulation time of 20 ms per window, was employed.

The acquired DIA-MS data was analyzed on Spectronaut Pulsar (v18.2.23, Biognosys, Schlieren, Zurich, Switzerland). A de novo spectral library was first prepared by searching the entire data set against the UniProtKB human protein database (UP000005640, 20,644 gene count) with default search parameters. The library comprised 8150 precursors and 5627 peptides corresponding to 1354 proteins. The raw files were then searched against the generated library and the human database using default settings: quantification at the MS2 level, differential abundance testing using an unpaired Student’s t-test, and global TIC (total intensity count) was used for normalization of data. FDR was set to 1% at both peptide and protein levels. The extracted areas under the peak of peptides were further used for quantitative analysis of the plasma proteome.

### MRM^HR^ acquisition and processing (Phase 1-Verification and Phase 2)

Plasma was digested with the same protocol as discussed in discovery study, resuspended in solvent A and spiked with 100 fmoles per injection of beta-galactosidase (β-gal, Sciex, MA, USA) as an internal standard. Targeted mass spectrometry was carried out in MRM^HR^ mode on the ZenoTOF 7600 system using the same trap and analytical column as mentioned above. For this, the y and b ions of the target proteins were selected from the SRMatlas Database^32^ as well as from the discovery Phase DIA-MS data. Initially, all the ions were scanned in an unscheduled MRM^HR^ mode on a pooled plasma sample to optimize the retention time (RT) of the ions. The product ions with a Gaussian peak shape and intensity of more than 100 counts per second (cps) were further selected and acquired in a scheduled MRM^HR^ mode on the pooled sample. Out of the initially selected 42 proteins, we could not successfully select any product ions/transitions for two proteins, namely RFX7 and IGHV3-20, based on the above-mentioned criteria. For the rest of the 40 proteins, a minimum of one peptide was selected, with at least two transitions per peptide for the majority. Finally, the optimized method consisting of 159 transitions (Table S4), with an accumulation time of 0.09s per transition, retention time tolerance of 33s and total run time of 50 min, was used to acquire data for all the plasma samples. The samples were eluted at a flow rate of 5 μL/min in a linear gradient of 5% solvent B (100% ACN (v/v) with 0.1% formic acid) to 80% solvent B in 45 min with a total run time of 50 min. The raw MRM^HR^ data was analyzed using the Analytics software integrated with Sciex OS platform (v 3.1.6.44, Sciex, MA, USA). The extracted peaks were selected based on expected RT, pre-processed by smoothing on a moving average, and filtered based on the following criteria: minimum peak height of 100 cps and minimum signal/noise ratio of 2. All selected spectral peaks were checked manually to ensure correct peak detection and accurate integration. The areas under the peak of selected product ions were used for all downstream processing.

### Data analysis and statistical methods

#### Phase 1-Discovery data analysis

The normalized intensities of the protein groups quantified in the samples were extracted from Spectronaut. Since plasma proteomics suffers from a higher percentage of missing values in comparison to cellular proteomics^33^, we removed all the proteins with excessive missing values (>40% in the entire data set). The remaining missing values were imputed using the K-nearest neighbor (KNN, K=5) method, employing the KNNImputer function from scikit-learn^34^. The imputed data were log_2_ transformed to generate a comprehensive downstream analysis dataset.

The processed data were visualized via principal component analysis (PCA), and the coefficient of variation (CV); median, and interquartile range were calculated to assess the quality of the acquired data. The proteomics data were evaluated using a two-tailed, unpaired Student’s t-test employing multiple testing correction. The candidates were filtered out by adjusted p-value (<0.05) and absolute log_2_ fold change of 0.58. The unique and overlapping DEPs identified in EMT and LMT samples were visualized using an Upset plot generated using the UpSetR package^35^ from the

Comprehensive R Archive Network (CRAN). Protein-protein interaction (PPI) map of DEPs was generated by NetworkAnalyst^36^ using a confidence score cutoff of 700 on the STRING interactome database. The functional modules were assigned using the InfoMap algorithm and were enriched for molecular functions using the GO: MF database. Gene set enrichment analysis (GSEA) was carried out using the clusterProfiler R package using the biological pathway (BP) as the gene ontology database^37^. We used the p-value cutoff of <0.05 to identify the pathways significantly altered in sPTB.

To complement our differential expression analysis, we used feature extraction algorithms to extract proteins important for the prediction of cases. Recursive feature elimination-Cross validation (RFECV, cv=5) from scikit-learn and SHapley Additive exPlannations^38^ (SHAP) employing three different classifiers viz. Support vector machine (SVM), Logistic regression (LR) and Random Forest (RF) were fitted on the entire discovery data of EMT samples to extract the proteins that theoretically have the highest likelihood of predicting sPTB. The selected proteins were then overlapped with the DEPs to obtain a comprehensive list common to both approaches. These proteins were subsequently verified for their expression pattern in the same sample set using the targeted MRM^HR^.

#### Verification (Phase 1) and Validation (Phase 2) data analysis

For the verification phase, firstly, the transitions with high missing values (in 15 or more samples) were removed; the remaining transitions were subjected to data pre-processing: log_2_ transformation, median centric normalization using the proBatch R package^39^, and missing data imputation employing the missForest package^40^. The imputed data were converted to multiples of medians, and the regulated transitions between sPTB and TB groups were statistically evaluated using an unpaired two-sided Wilcoxon rank sum test from the R stats package (v3.6.2). Due to a limited number of data points, no multiple testing corrections were performed in the verification targeted analysis^41^.

Next, all the statistically significant transitions that were regulated in the opposite direction from their corresponding discovery phase protein data were removed from downstream analysis. We used the Boruta feature extraction method^42^(maxRuns=100) to identify the transitions having predictive power, which can be selected for validation in a separate sample set and for the development of a prediction model. Confirmed transitions from Boruta were overlapped with transitions significant on Wilcoxon rank sum test; peptides corresponding to the common transitions were measured in Phase 2 plasma samples.

Peptide expression levels measured in the validation sample set were pre-processed using the same pipeline as in the verification data. Next, we attempted to build prediction models for predicting the risk of sPTB. The schematic for the development and testing of models is represented in Figure 6B. We selected 4 different binary classifiers, namely SVM, LR, RF and XGBoost (XGB), based on their level of complexity. All the classifiers were trained using the sample set from the verification arm and were tested on samples from Phase 2. Hyperparameter tuning of all classifiers was done using GridSearchCV and RandomizedSearchCV from sklearn.model selection. We developed different models based on a combination of candidate protein markers as well as the clinical and demographic parameters of the mothers. All the feature combinations were fitted into the models using every binary classifier algorithm selected. For model 1, we combined the plasma expression levels of *MICAL2* and *CAPS2*, whereas for model 2, we trained and tested the model only for the *CAPS2* marker.

For developing the models using the clinical and demographic parameters of the mothers, first, we selected the 33 relevant and available parameters from the Garbh-Ini cohort. Thereafter, we employed the RFE-CV (CV=5) algorithm to extract the most predictive variables out of them. The selected parameters were then used to build model 3 and were tested for their predictive power of sPTB. *CAPS2* expression levels were later combined with these RFE-CV selected features as a part of model 4 to assess the power of a comprehensive prediction algorithm. All the models were evaluated using area under the Receiver operating curve (AUROC), area under the Precision-Recall curve (AUPRC). Models’ accuracy negative predictive value (NPV), positive predictive value (PPV), sensitivity and specificity were calculated using sklearn.metrics across a range of prediction probability thresholds and was visualized with a threshold performance graph plotted using the runway package from RStudio. Bootstrapping (n_iterations=1000) was calculated using a resample function from sklearn.utils to calculate the 95% confidence intervals for all the metrics.

## Acknowledgement

We sincerely thank the research physicians, study nurses, lab technicians, field workers, Internal Quality Improvement team, project members, and data management staff for their dedication and contributions. Special appreciation goes to the RCB mass spectrometry facility for its ongoing support. GARBH-Ini program (Interdisciplinary Group for Advanced Research on Birth Outcomes), an initiative by the Department of Biotechnology (DBT), Government of India, is coordinated by THSTI. This program unites expertise from multiple research institutions like THSTI, the National Institute of Biomedical Genomics (NIBMG) in Kalyani, and the Regional Centre for Biotechnology (RCB) in the National Capital Region, in collaboration with healthcare centers, including Gurugram Civil Hospital (GCH) and major New Delhi hospitals like Safdarjung Hospital and Maulana Azad Medical College (MAMC). The initiative focuses on providing essential tools to clinicians and policymakers by developing (i) risk stratification algorithms for preterm birth (PTB) using clinical and demographic data, (ii) biomarkers from simple biochemical to complex omics markers for personalized decision-making, and (iii) predictive models for health systems at both individual and population levels. To achieve these goals, the program uses precise ultrasonographic pregnancy dating and local data-driven AI models to identify cases of fetal growth restriction and preterm birth. This improves the accuracy of predictive algorithms and enhances understanding of potential biological causes. As part of its efforts, the GARBH-Ini pregnancy cohort, established in May 2015, enrolls women before 20 weeks of pregnancy and monitors them throughout, collecting comprehensive clinical data, biospecimens and conducting serial ultrasounds. Detailed information is available at https://www.garbhinicohort.in.

## Author Contributions

N.K., A.B., S.Bai, and S.S. did the sample preparation and method development for phase 1. N.K. and A.B. performed phase 1 data acquisition and data analysis. N.K. and S.S.H. prepared samples for phase 2, and N.K. did method development, data acquisition and analysis for phase 2. N.S. and B.K.D. supported in building AI/ML models. P.K. provided substantial contributions to sample processing and biorepository management. N.W. was involved in clinical study monitoring and data management. R.T. substantially contributed to clinical analysis, study design and data analysis. D.M.S. contributed to the Study design and manuscript writing. S.B. established the cohort, coordinated and designed the study. T.K.M. substantially contributed to the study design, coordination of the proteomics study, development of methods, and data analysis. N.K., T.K.M., R.T., S.B, D.M.S. and A.T. wrote the manuscript with input and substantial revisions from all the authors.

## GARBH-Ini study group’s contribution

Shinjini Bhatnagar (S.B.), Nitya Wadhwa (N.W.), Uma Chandra Mouli Natchu (U.C.M.N.), Ramachandran Thiruvengadam ( R.T.), Sumit Misra (S.M.), Dharmendra Sharma (D.S.), Kanika Sachdeva (K.S.), Amanpreet Singh (A.S.), Satyajit Rath (S.R.), and Vineeta Bal(V.B.) Alka Sharma (A.S.), Sunita Sharma(S.S.), Umesh Mehta (U.M.), and Brahmdeep Sindhu (B.S.) Pratima Mittal (P.M.), Rekha Bharti (R.B.), Harish Chellani (H.C.), Rani Gera (R.G.), Jyotsna Suri(J.S.), Pradeep Debata(P.D.), and Sugandha Arya(S.A.), Nikhil Tandon (N.T.), Yashdeep Gupta (Y.G), Alpesh Goyal (A.G.), Smriti Hari(S.H.), Aparna Sharma K (A.S.K.), Anubhuti Rana (A.R.), Rakesh Gupta ( R.G.) Siddarth Ramji (S.R.) and Anju Garg (A.G.), Ashok Khurana (A.K.), Reva Tripathi ( R.T.), Himanshu Sinha (H.S.), and Raghunathan Rengasamy (R.R) contributed to the clinical study design, data, and biospecimen collection, and quality assurance. Tushar K Maiti (T.K.M), Arindam Maitra (A.M.), Bhabatosh Das (B.D.), Pallavi S Kshetrapal (P.S.K.), Shailaja Sopory (S.S.), Balakrish G Nair (G.B.N), Dinakar M Salunke (D.M.S), and Partha P Majumder (P.P.M) contributed to the design of the study, supervision of laboratory experiments, and biochemical analysis.

## Ethics approval

The study was approved by the Institutional Ethics Committee of all participating Institutions, Translational Health Science and Technology Institute, and the Regional Centre for Biotechnology.

## Data availability

The mass spectrometric DIA data generated for the discovery study have been deposited in the ProteomeXchange database under accession code PXD064010, whereas the MRM^HR^ data for verification and validation are deposited under accession code PXD065087.

