## Supplementary Information for "Plasma proteome profiling identifies predictive signatures for preterm birth risk"

Naman Kharbanda *et al.*

*** Corresponding authors:**

Dr. Tushar Kanti Maiti,

Dr. Shinjini Bhatnagar

Dr. Dinakar M Salunke

**This PDF file includes:**

Figs. S1 to S5
Tables S1 to S9

**Supplementary Figures:**

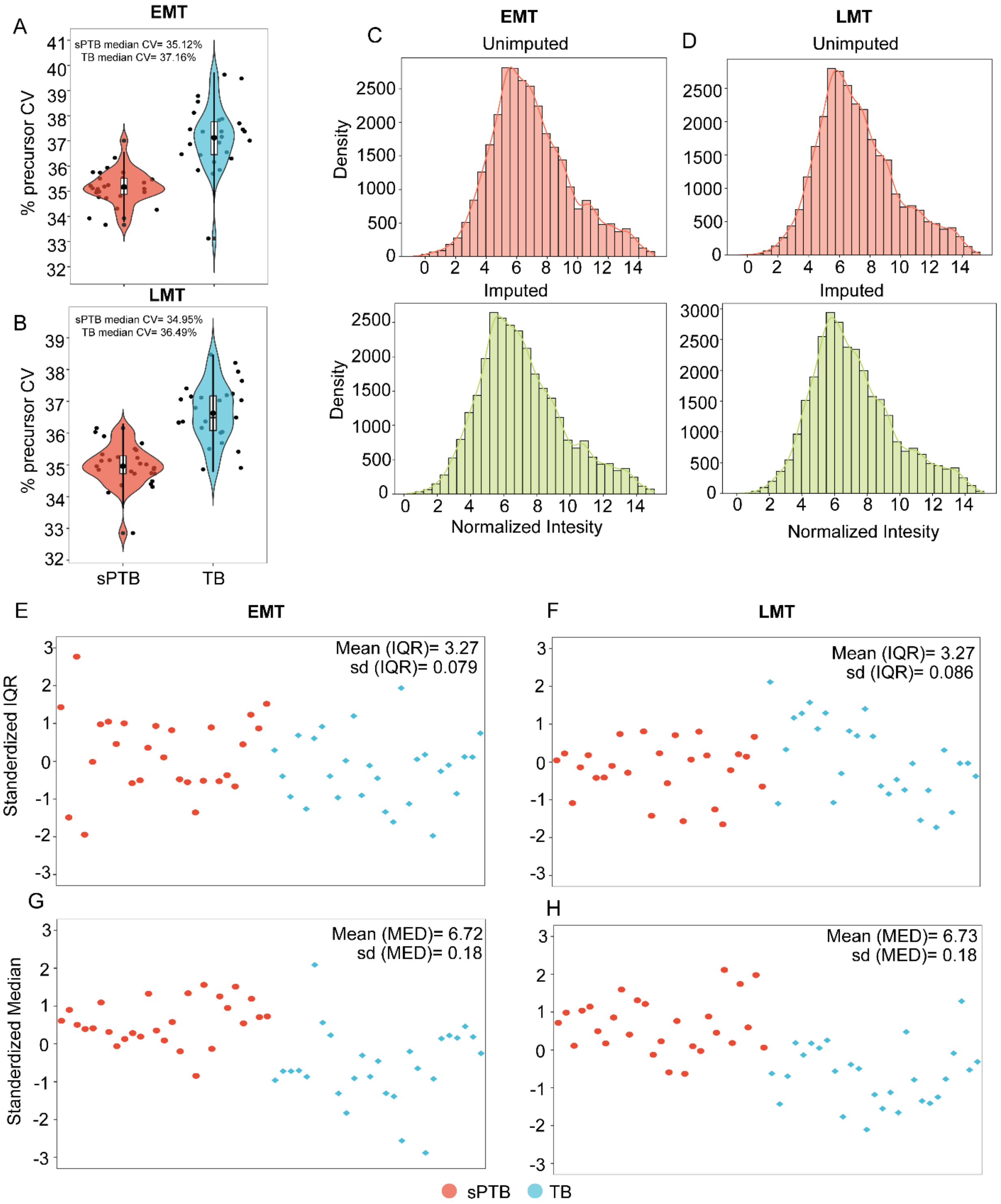


**Fig. S1: Quality control for discovery proteomics data acquisition. (A, B)** Intragroup precursor % coefficient of variation displayed in a violin plot overlaid with a box and whisker plot and a scatter plot denoting variation in the individual samples. **(C, D)** Intensity distribution of normalized data pre and post imputation, showcasing the Gaussian spread in both EMT and LMT groups. **(E, F)** Standardized IQR of protein expressions in the sPTB and TB samples. **(G, H)** Median protein expression in individual plasma samples.

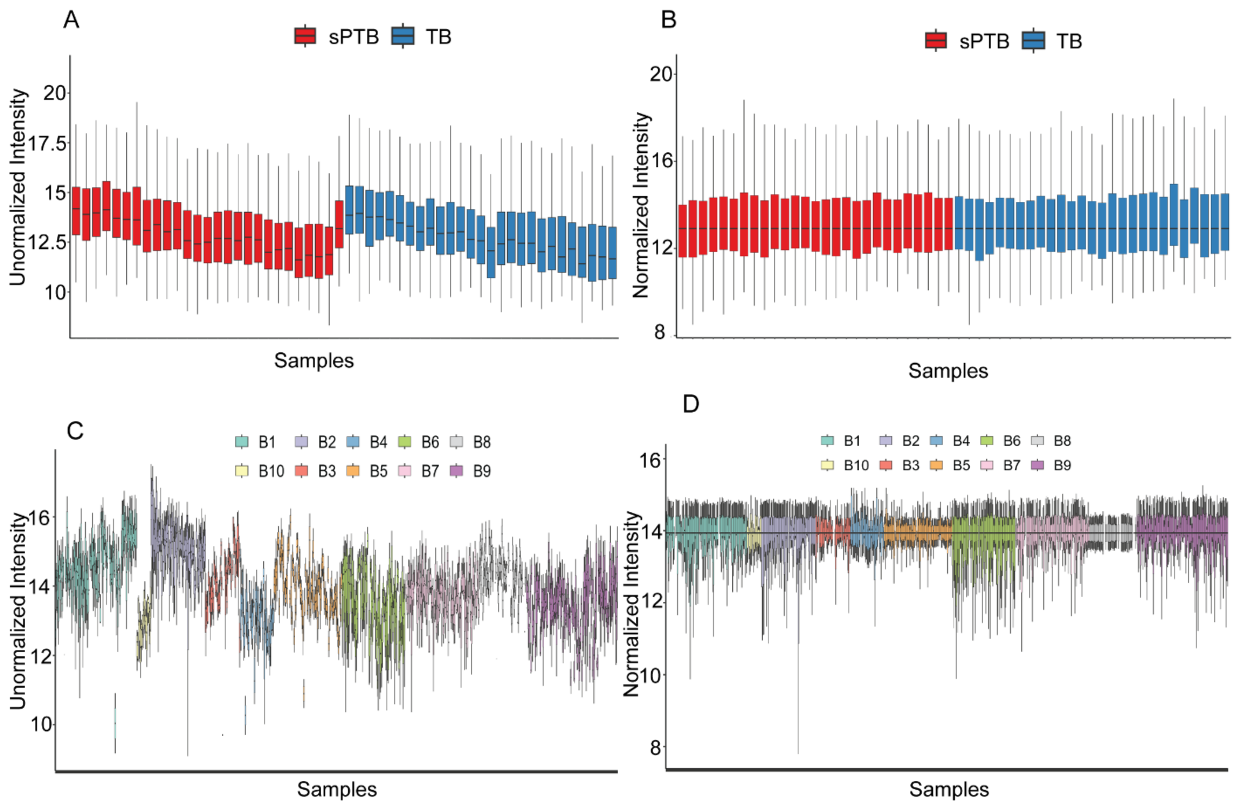


**Fig. S2: Median centric normalization in Phase 1 (verification) and Phase 2. (A, B)** Unnormalized and normalized response of unimputed MRM$^{HR}$ data collected in Phase 1 (verification) (n=54). **(C, D)** Unnormalized and normalized response of unimputed MRM$^{HR}$ data collected in different batches in Phase 2 (n=795).

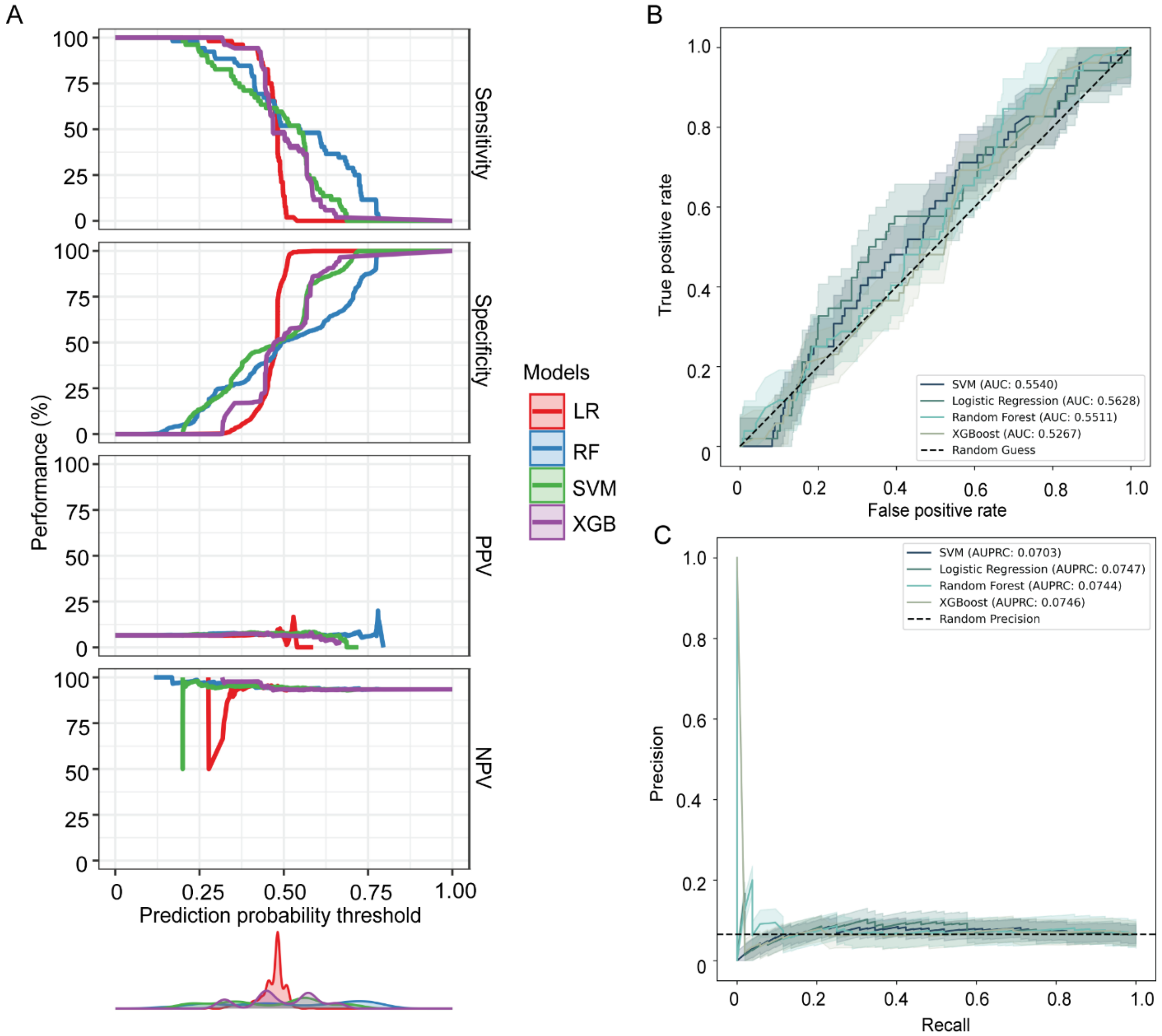


**Fig. S3: Performance of the prediction model 1 developed using *CAPS2* and *MICAL2* levels. (A)** Prediction performance of model 1 developed using different classifiers depicted by sensitivity, specificity, PPV and NPV calculated at multiple prediction probability cutoff. **(B, C)** Receivers operating curve and precision recall curve displaying the prediction performance of individual models. AUROC and AUPRC are mentioned in the lower right and upper right of the plots respectively. The shaded region represents the 95% CI. PPV: Positive predictive value, NPV: Negative predictive value

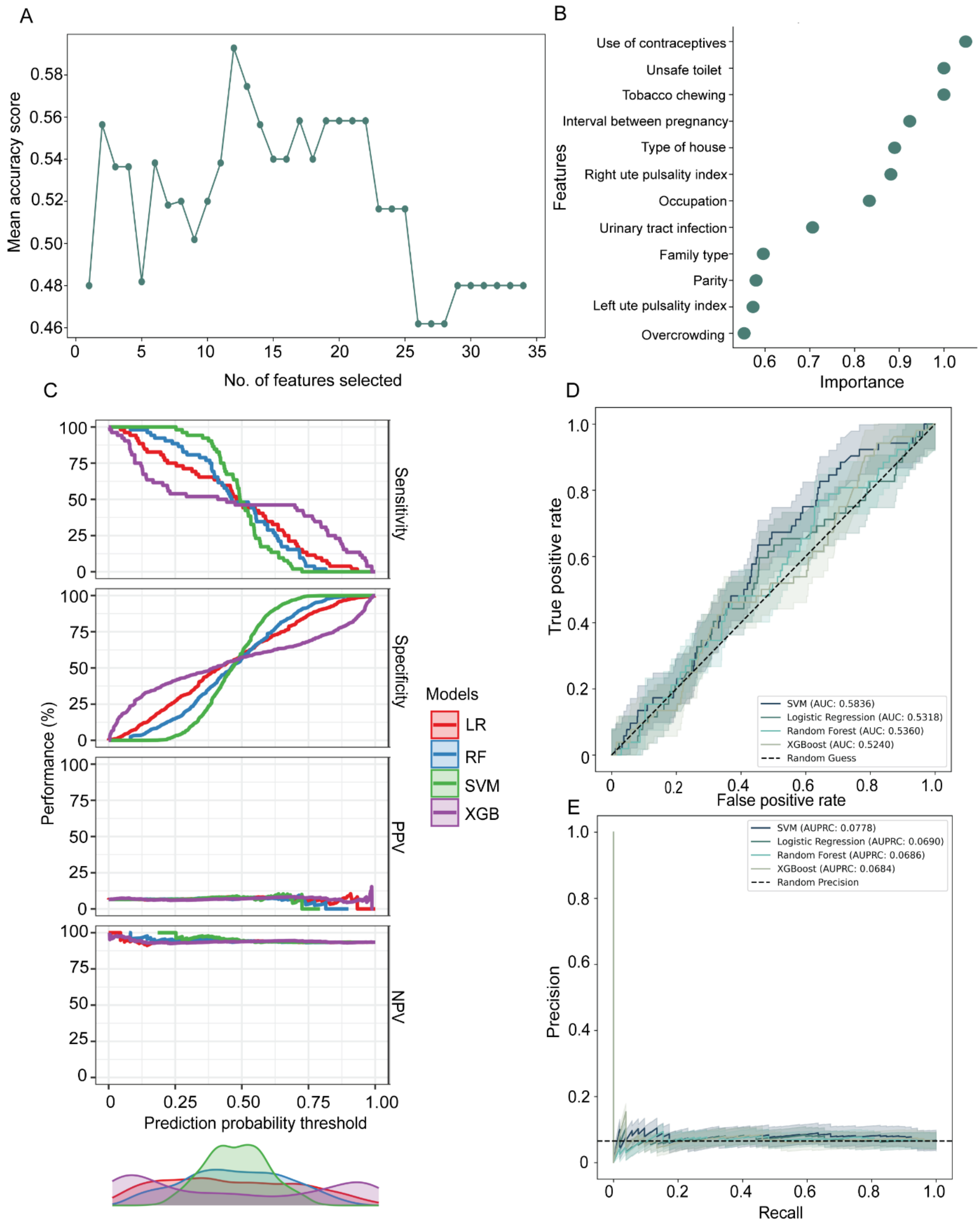


**Fig. S4: Prediction performance of model 3 developed using clinical and demographic parameters. (A)** Mean accuracy score calculated over 5-fold by RFE-CV for different clinical and demographic feature subsets. **(B)** RFE-CV selected features sorted based on their feature importance score. **(C)** Prediction performance of model 3 developed using different classifiers depicted by sensitivity, specificity, PPV and NPV calculated at multiple prediction probability cutoff. **(D, E)** Receivers operating curve and precision recall curve displaying the prediction performance of individual models. AUROC and AUPRC are mentioned in the lower right and upper right of the plots respectively. The shaded region represents the 95% CI. PPV: Positive predictive value, NPV: Negative predictive value

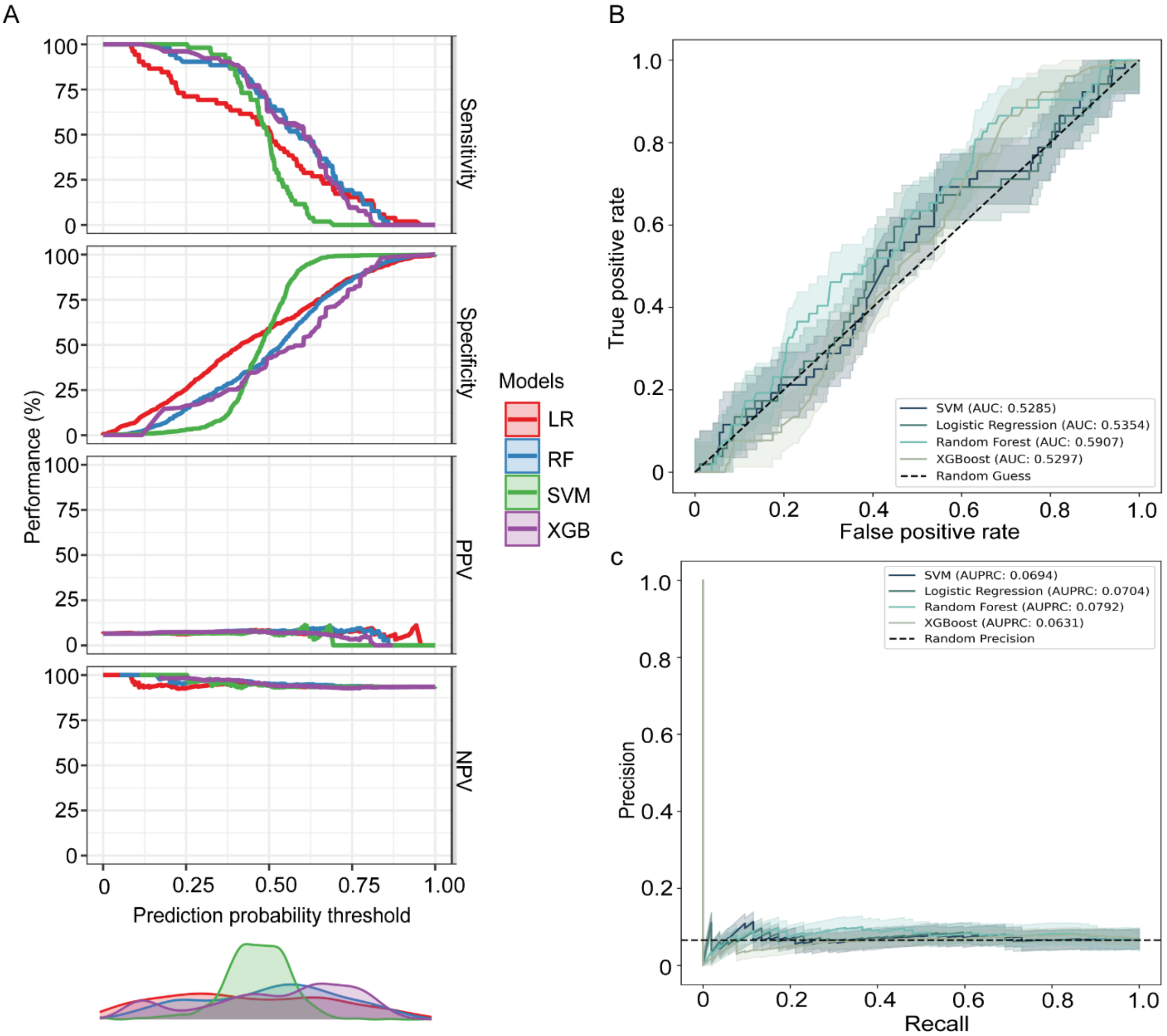


**Fig. S5: Comprehensive model combining clinical and demographic parameters with protein marker. (A)** Prediction performance of model 4 developed using different classifiers depicted by sensitivity, specificity, PPV and NPV calculated at multiple prediction probability cutoff. **(B, C)** Combination of clinical and demographic parameters with *CAPS2* expression did not show significant improvement in prediction on both receivers operating curve and precision recall curve. AUROC and AUPRC are mentioned in the lower right and upper right of the plots respectively. PPV: Positive predictive value, NPV: Negative predictive value.

### Supplementary Tables:

**Table S1:** Differentially expressed proteins between sPTB and TB groups in plasma samples collected at the EMT stage.

| S. No. | Uniprot ID | Gene name | Protein description | Fold change[a] (sPTB/TB) | p-value[b] | q-value[c] |
|---|---|---|---|---|---|---|
| 1 | Q8N9V7 | *TOPAZ1* | Protein TOPAZ1 | 2.00 | 3E-84 | 4E-82 |
| 2 | Q7Z3Z0 | *KRT25* | Keratin, type I cytoskeletal 25 | 2.27 | 4E-72 | 2E-70 |
| 3 | Q86SR1 | *GALNT10* | Polypeptide N-acetylgalactosaminyltransferase 10 | 0.59 | 1E-71 | 5E-70 |
| 4 | Q4V328 | *GRIPAP1* | GRIP1-associated protein 1 | 2.23 | 1E-59 | 2E-58 |
| 5 | A6NE01 | *FAM186A* | Protein FAM186A | 2.67 | 1E-55 | 2E-54 |
| 6 | Q92782 | *DPF1* | Zinc finger protein neuro-d4 | 2.98 | 8E-50 | 1E-48 |
| 7 | Q86UP8 | *GTF2IRD2* | General transcription factor II-I repeat domain-containing protein 2A | 2.78 | 3E-49 | 3E-48 |
| 8 | Q9NQS7 | *INCENP* | Inner centromere protein | 1.78 | 5E-47 | 4E-46 |
| 9 | Q2KHR2 | *RFX7* | DNA-binding protein RFX7 | 0.09 | 7E-47 | 6E-46 |
| 10 | P13569 | *CFTR* | Cystic fibrosis transmembrane conductance regulator | 1.73 | 1E-45 | 9E-45 |
| 11 | Q7Z7G8 | *VPS13B* | Vacuolar protein sorting-associated protein 13B | 1.78 | 2E-44 | 1E-43 |
| 12 | Q4G148 | *GXYLT1* | Glucoside xylosyltransferase 1 | 2.87 | 5E-43 | 3E-42 |
| 13 | Q9C0C9 | *UBE2O* | (E3-independent) E2 ubiquitin-conjugating enzyme | 1.90 | 2E-40 | 1E-39 |
| 14 | Q8N9U0 | *TC2N* | Tandem C2 domains nuclear protein | 1.90 | 2E-39 | 7E-39 |
| 15 | Q9NR34 | *MAN1C1* | Mannosyl-oligosaccharide 1,2-alpha-mannosidase IC | 1.52 | 9E-38 | 3E-37 |
| 16 | Q9BXY5 | *CAPS2* | Calcyphosin-2 | 2.21 | 2E-37 | 7E-37 |
| 17 | P01709 | *IGLV2-8* | Immunoglobulin lambda variable 2-8 | 0.50 | 3E-36 | 9E-36 |
| 18 | Q14376 | *GALE* | UDP-glucose 4-epimerase | 1.55 | 5E-36 | 2E-35 |
| 19 | Q96K80 | *ZC3H10* | Zinc finger CCCH domain-containing protein 10 | 3.07 | 8E-36 | 2E-35 |
| 20 | Q99459 | *CDC5L* | Cell division cycle 5-like protein | 2.14 | 4E-34 | 1E-33 |
| 21 | Q05215 | *EGR4* | Early growth response protein 4 | 1.88 | 4E-34 | 1E-33 |
| 22 | Q68D10 | *SPTY2D1* | Protein SPT2 homolog | 2.70 | 2E-33 | 5E-33 |
| 23 | P58107 | *EPPK1* | Epiplakin | 1.89 | 2E-33 | 6E-33 |
| 24 | Q5JRA6 | *MIA3* | Transport and Golgi organization protein 1 homolog | 1.96 | 4E-33 | 8E-33 |
| 25 | O43790 | *KRT86* | Keratin, type II cuticular Hb6 | 0.66 | 1E-31 | 2E-31 |
| 26 | Q9NQ55 | *PPAN* | Suppressor of SWI4 1 homolog | 0.52 | 1E-30 | 2E-30 |
| 27 | Q96JE9 | *MAP6* | Microtubule-associated protein 6 | 2.37 | 3E-29 | 5E-29 |
| 28 | P19838 | *NFKB1* | Nuclear factor NF-kappa-B p105 subunit | 2.69 | 3E-29 | 5E-29 |
| 29 | P01817 | *IGHV2-5* | Immunoglobulin heavy variable 2-5 | 1.87 | 2E-26 | 3E-26 |
| 30 | P01185 | *AVP* | Vasopressin-neurophysin 2-copeptin | 1.66 | 3E-26 | 5E-26 |
| 31 | P22455 | *FGFR4* | Fibroblast growth factor receptor 4 | 2.44 | 1E-25 | 2E-25 |
| 32 | Q92543 | *SNX19* | Sorting nexin-19 | 1.71 | 3E-25 | 5E-25 |
| 33 | Q8TDI8 | *TMC1* | Transmembrane channel-like protein 1 | 2.38 | 2E-24 | 3E-24 |
| 34 | Q6MZP7 | *LIN54* | Protein lin-54 homolog | 2.94 | 4E-24 | 6E-24 |
| 35 | P34820 | *BMP8B* | Bone morphogenetic protein 8B | 0.47 | 1E-21 | 1E-21 |
| 36 | A0A0J9YXX1 | *IGHV5-10-1* | Immunoglobulin heavy variable 5-10-1 | 0.49 | 4E-21 | 5E-21 |
| 37 | P69905 | *HBA1* | Hemoglobin subunit alpha | 0.65 | 9E-20 | 1E-19 |
| 38 | Q5T200 | *ZC3H13* | Zinc finger CCCH domain-containing protein 13 | 1.78 | 9E-19 | 1E-18 |
| 39 | P42679 | *MATK* | Megakaryocyte-associated tyrosine-protein kinase | 2.04 | 4E-18 | 5E-18 |
| 40 | Q92952 | *KCNN1* | Small conductance calcium-activated potassium channel protein 1 | 1.52 | 5E-18 | 6E-18 |
| 41 | Q59EK9 | *RUNDC3A* | RUN domain-containing protein 3A | 2.08 | 7E-17 | 7E-17 |
| 42 | Q9Y4G2 | *PLEKHM1* | Pleckstrin homology domain-containing family M member 1 | 2.33 | 2E-16 | 2E-16 |
| 43 | O94851 | *MICAL2* | [F-actin]-monooxygenase MICAL2 | 1.81 | 3E-16 | 3E-16 |
| 44 | Q9H9J4 | *USP42* | Ubiquitin carboxyl-terminal hydrolase 42 | 1.57 | 5E-16 | 5E-16 |
| 45 | Q6Y7W6 | *GIGYF2* | GRB10-interacting GYF protein 2 | 1.96 | 2E-15 | 2E-15 |
| 46 | Q008S8 | *ECT2L* | Epithelial cell-transforming sequence 2 oncogene-like | 1.63 | 3E-14 | 2E-14 |
| 47 | Q9NT99 | *LRRC4B* | Leucine-rich repeat-containing protein 4B | 1.61 | 4E-14 | 3E-14 |
| 48 | Q14966 | *ZNF638* | Zinc finger protein 638 | 1.58 | 1E-13 | 1E-13 |
| 49 | O00160 | *MYO1F* | Unconventional myosin-If | 1.86 | 1E-13 | 1E-13 |

| | | | | | | |
|---|---|---|---|---|---|---|
| 50 | Q6XR72 | *SLC30A10* | Zinc transporter 10 | 1.70 | 2E-13 | 2E-13 |
| 51 | P25940 | *COL5A3* | Collagen alpha-3(V) chain | 1.62 | 5E-13 | 4E-13 |
| 52 | Q14643 | *ITPR1* | Inositol 1,4,5-trisphosphate receptor type 1 | 1.78 | 1E-12 | 8E-13 |
| 53 | Q8WWB5 | *PIH1D2* | PIH1 domain-containing protein 2 | 1.56 | 1E-12 | 1E-12 |
| 54 | Q13451 | *FKBP5* | Peptidyl-prolyl cis-trans isomerase FKBP5 | 1.75 | 2E-12 | 2E-12 |
| 55 | Q15465 | *SHH* | Sonic hedgehog protein | 0.58 | 4E-12 | 3E-12 |
| 56 | P82094 | *TMF1* | TATA element modulatory factor | 1.74 | 7E-12 | 5E-12 |
| 57 | P01593 | *IGKV1D-33* | Immunoglobulin kappa variable 1D-33 | 1.70 | 7E-11 | 5E-11 |
| 58 | A0A0C4DH32 | *IGHV3-20* | Immunoglobulin heavy variable 3-20 | 0.42 | 8E-11 | 5E-11 |
| 59 | O75592 | *MYCBP2* | E3 ubiquitin-protein ligase MYCBP2 | 2.08 | 3E-10 | 2E-10 |
| 60 | O95696 | *BRD1* | Bromodomain-containing protein 1 | 1.98 | 4E-10 | 2E-10 |
| 61 | A4D126 | *CRPPA* | D-ribitol-5-phosphate cytidylyltransferase | 1.71 | 2E-09 | 1E-09 |
| 62 | P09919 | *CSF3* | Granulocyte colony-stimulating factor | 1.72 | 4E-09 | 2E-09 |
| 63 | Q96I82 | *KAZALD1* | Kazal-type serine protease inhibitor domain-containing protein 1 | 1.68 | 9E-09 | 5E-09 |
| 64 | Q9UPS6 | *SETD1B* | Histone-lysine N-methyltransferase SETD1B | 2.14 | 1E-08 | 7E-09 |
| 65 | P09619 | *PDGFRB* | Platelet-derived growth factor receptor beta | 1.66 | 2E-08 | 9E-09 |
| 66 | A0A075B6I0 | *IGLV8-61* | Immunoglobulin lambda variable 8-61 | 1.51 | 3E-08 | 2E-08 |
| 67 | P29590 | *PML* | Protein PML | 1.55 | 5E-08 | 3E-08 |
| 68 | Q9NU22 | *MDN1* | Midasin | 1.79 | 9E-08 | 5E-08 |
| 69 | Q9NQU5 | *PAK6* | Serine/threonine-protein kinase PAK 6 | 1.54 | 2E-07 | 1E-07 |
| 70 | P05109 | *S100A8* | Protein S100-A8 | 2.67 | 3E-07 | 1E-07 |
| 71 | Q6VAB6 | *KSR2* | Kinase suppressor of Ras 2 | 1.78 | 3E-07 | 2E-07 |
| 72 | P51149 | *RAB7A* | Ras-related protein Rab-7a | 1.68 | 3E-07 | 2E-07 |
| 73 | P45880 | *VDAC2* | Voltage-dependent anion-selective channel protein 2 | 0.56 | 4E-07 | 2E-07 |
| 74 | Q96SA4 | *SERINC2* | Serine incorporator 2 | 1.54 | 4E-07 | 2E-07 |
| 75 | P08294 | *SOD3* | Extracellular superoxide dismutase [Cu-Zn] | 0.54 | 4E-07 | 2E-07 |
| 76 | P0DJI8 | *SAA1* | Serum amyloid A-1 protein | 3.89 | 5E-07 | 2E-07 |
| 77 | P57737 | *CORO7* | Coronin-7 | 1.60 | 2E-06 | 1E-06 |
| 78 | A6NDX5 | *ZNF840P* | Putative zinc finger protein 840 | 1.76 | 3E-06 | 2E-06 |
| 79 | Q9BV68 | *RNF126* | E3 ubiquitin-protein ligase RNF126 | 1.52 | 7E-06 | 3E-06 |
| 80 | Q8NCW5 | *NAXE* | NAD(P)H-hydrate epimerase | 1.92 | 1E-05 | 4E-06 |
| 81 | Q5SYB0 | *FRMPD1* | FERM and PDZ domain-containing protein 1 | 1.77 | 1E-05 | 5E-06 |
| 82 | Q9UQ72 | *PSG11* | Pregnancy-specific beta-1-glycoprotein 11 | 1.97 | 6E-05 | 2E-05 |
| 83 | Q8IV61 | *RASGRP3* | Ras guanyl-releasing protein 3 | 0.64 | 8E-05 | 3E-05 |
| 84 | P0CG23 | *ZNF853* | Zinc finger protein 853 | 2.47 | 2E-04 | 7E-05 |
| 85 | Q9H116 | *GZF1* | GDNF-inducible zinc finger protein 1 | 0.60 | 2E-04 | 9E-05 |
| 86 | A6ND36 | *FAM83G* | Protein FAM83G | 2.21 | 4E-04 | 1E-04 |
| 87 | P20073 | *ANXA7* | Annexin A7 | 1.66 | 5E-04 | 2E-04 |
| 88 | P20718 | *GZMH* | Granzyme H | 1.86 | 7E-04 | 2E-04 |
| 89 | Q9P2S5 | *WRAP73* | WD repeat-containing protein WRAP73 | 0.37 | 1E-03 | 4E-04 |
| 90 | Q96D09 | *GPRASP2* | G-protein coupled receptor-associated sorting protein 2 | 1.68 | 2E-03 | 5E-04 |
| 91 | Q04695 | *KRT17* | Keratin, type I cytoskeletal 17 | 1.53 | 2E-03 | 5E-04 |
| 92 | P02741 | *CRP* | C-reactive protein | 1.57 | 2E-03 | 7E-04 |
| 93 | P00709 | *LALBA* | Alpha-lactalbumin | 1.87 | 2E-03 | 8E-04 |
| 94 | Q96AC6 | *KIFC2* | Kinesin-like protein KIFC2 | 1.52 | 2E-03 | 8E-04 |
| 95 | Q96PE1 | *ADGRA2* | Adhesion G protein-coupled receptor A2 | 1.85 | 3E-03 | 9E-04 |
| 96 | Q8IWY4 | *SCUBE1* | Signal peptide, CUB and EGF-like domain-containing protein 1 | 0.59 | 3E-03 | 1E-03 |
| 97 | P02533 | *KRT14* | Keratin, type I cytoskeletal 14 | 1.50 | 5E-03 | 1E-03 |
| 98 | A0A2R8Y619 | *H2BK1* | Histone H2B type 2-K1 | 8.59 | 2E-02 | 4E-03 |
| 99 | Q9Y5J7 | *TIMM9* | Mitochondrial import inner membrane translocase subunit Tim9 | 0.50 | 2E-02 | 5E-03 |
| 100 | Q96L34 | *MARK4* | MAP/microtubule affinity-regulating kinase 4 | 2.00 | 2E-02 | 5E-03 |
| 101 | Q15554 | *TERF2* | Telomeric repeat-binding factor 2 | 0.42 | 2E-02 | 5E-03 |
| 102 | P08670 | *VIM* | Vimentin | 2.52 | 2E-02 | 6E-03 |
| 103 | Q9UM73 | *ALK* | ALK tyrosine kinase receptor | 0.24 | 2E-02 | 6E-03 |
| 104 | P24821 | *TNC* | Tenascin | 0.38 | 3E-02 | 8E-03 |
| 105 | Q9NZU7 | *CABP1* | Calcium-binding protein 1 | 0.37 | 4E-02 | 9E-03 |
| 106 | Q15063 | *POSTN* | Periostin | 1.97 | 4E-02 | 1E-02 |
| 107 | Q15643 | *TRIP11* | Thyroid receptor-interacting protein 11 | 0.18 | 5E-02 | 1E-02 |

| | | | | | | |
|---|---|---|---|---|---|---|
| 108 | Q9Y4D2 | *DAGLA* | Diacylglycerol lipase-alpha | 1.68 | 5E-02 | 1E-02 |
| 109 | P37840 | *SNCA* | Alpha-synuclein | 1.54 | 7E-02 | 2E-02 |
| 110 | Q6NT04 | *TIGD7* | Tigger transposable element-derived protein 7 | 1.80 | 7E-02 | 2E-02 |
| 111 | O95803 | *NDST3* | Bifunctional heparan sulfate | 1.79 | 8E-02 | 2E-02 |
| 112 | Q9NSG2 | *C1orf112* | Uncharacterized protein C1orf112 | 1.54 | 9E-02 | 2E-02 |
| 113 | P0DJI9 | *SAA2* | Serum amyloid A-2 protein | 2.25 | 1E-01 | 2E-02 |

[a]Fold change cutoff: Upregulation ≥1.5, Downregulation ≤0.66. Unpaired student's t-test with FDR correction was used for statistical evaluation in Spectronaut. [b] p-value <0.05 was considered statistically significant. [c] FDR-adjusted p-value (q-value) <0.05 was considered statistically significant. sPTB: Spontaneous preterm birth, TB: Term birth, EMT: Early-mid trimester.

\

**Table S2:** Differentially expressed proteins between sPTB and TB groups in plasma samples collected at LMT stage.

| S. NO. | Uniprot ID | Gene Name | Protein description | Fold change[a] (sPTB/TB) | p-value[b] | q-value[c] |
|---|---|---|---|---|---|---|
| 1 | Q8N9V7 | *TOPAZ1* | Protein TOPAZ1 | 1.84 | 1E-88 | 2E-86 |
| 2 | Q7Z3Z0 | *KRT25* | Keratin, type I cytoskeletal 25 | 2.12 | 7E-86 | 4E-84 |
| 3 | Q9BXY5 | *CAPS2* | Calcyphosin-2 | 2.00 | 6E-66 | 3E-64 |
| 4 | Q9NR34 | *MAN1C1* | Mannosyl-oligosaccharide 1,2-alpha-mannosidase IC | 1.82 | 3E-65 | 1E-63 |
| 5 | Q86SR1 | *GALNT10* | Polypeptide N-acetylgalactosaminyltransferase 10 | 0.60 | 7E-65 | 2E-63 |
| 6 | A6NE01 | *FAM186A* | Protein FAM186A | 2.49 | 2E-62 | 5E-61 |
| 7 | Q9UPS6 | *SETD1B* | Histone-lysine N-methyltransferase SETD1B | 2.75 | 3E-53 | 4E-52 |
| 8 | Q14376 | *GALE* | UDP-glucose 4-epimerase | 1.92 | 4E-52 | 4E-51 |
| 9 | P58107 | *EPPK1* | Epiplakin | 1.89 | 4E-52 | 4E-51 |
| 10 | Q4V328 | *GRIPAP1* | GRIP1-associated protein 1 | 2.06 | 3E-50 | 3E-49 |
| 11 | P13569 | *CFTR* | Cystic fibrosis transmembrane conductance regulator | 1.95 | 1E-49 | 9E-49 |
| 12 | Q9C0C9 | *UBE2O* | (E3-independent) E2 ubiquitin-conjugating enzyme | 2.11 | 4E-49 | 3E-48 |
| 13 | Q86UP8 | *GTF2IRD2* | General transcription factor II-I repeat domain-containing protein 2A | 2.48 | 4E-48 | 3E-47 |
| 14 | Q9Y4G2 | *PLEKHM1* | Pleckstrin homology domain-containing family M member 1 | 2.56 | 5E-48 | 3E-47 |
| 15 | Q9NQS7 | *INCENP* | Inner centromere protein | 1.97 | 2E-47 | 1E-46 |
| 16 | Q96K80 | *ZC3H10* | Zinc finger CCCH domain-containing protein 10 | 2.52 | 2E-46 | 1E-45 |
| 17 | Q99459 | *CDC5L* | Cell division cycle 5-like protein | 2.34 | 1E-45 | 6E-45 |
| 18 | Q68D10 | *SPTY2D1* | Protein SPT2 homolog | 2.35 | 1E-44 | 7E-44 |
| 19 | Q2KHR2 | *RFX7* | DNA-binding protein RFX7 | 0.05 | 6E-44 | 3E-43 |
| 20 | Q8N9U0 | *TC2N* | Tandem C2 domains nuclear protein | 1.99 | 2E-43 | 1E-42 |
| 21 | Q9NQ55 | *PPAN* | Suppressor of SWI4 1 homolog | 0.51 | 9E-40 | 4E-39 |
| 22 | Q92782 | *DPF1* | Zinc finger protein neuro-d4 | 2.45 | 4E-39 | 1E-38 |
| 23 | Q92543 | *SNX19* | Sorting nexin-19 | 1.57 | 4E-37 | 1E-36 |
| 24 | Q96JE9 | *MAP6* | Microtubule-associated protein 6 | 1.90 | 1E-36 | 3E-36 |
| 25 | P82094 | *TMF1* | TATA element modulatory factor | 1.67 | 1E-35 | 3E-35 |
| 26 | P42679 | *MATK* | Megakaryocyte-associated tyrosine-protein kinase | 1.83 | 7E-32 | 2E-31 |
| 27 | O75592 | *MYCBP2* | E3 ubiquitin-protein ligase MYCBP2 | 2.08 | 7E-32 | 2E-31 |
| 28 | Q5JRA6 | *MIA3* | Transport and Golgi organization protein 1 homolog | 1.69 | 3E-31 | 6E-31 |
| 29 | Q4G148 | *GXYLT1* | Glucoside xylosyltransferase 1 | 2.21 | 4E-30 | 9E-30 |
| 30 | Q6MZP7 | *LIN54* | Protein lin-54 homolog | 3.14 | 2E-29 | 3E-29 |
| 31 | O00160 | *MYO1F* | Unconventional myosin-If | 1.62 | 1E-28 | 2E-28 |
| 32 | Q05215 | *EGR4* | Early growth response protein 4 | 1.83 | 2E-28 | 3E-28 |
| 33 | Q8TDI8 | *TMC1* | Transmembrane channel-like protein 1 | 1.89 | 6E-28 | 1E-27 |
| 34 | P51149 | *RAB7A* | Ras-related protein Rab-7a | 1.97 | 3E-27 | 5E-27 |
| 35 | Q9NWZ5 | *UCKL1* | Uridine-cytidine kinase-like 1 | 0.65 | 1E-26 | 2E-26 |
| 36 | Q59EK9 | *RUNDC3A* | RUN domain-containing protein 3A | 2.09 | 2E-25 | 3E-25 |
| 37 | O94874 | *UFL1* | E3 UFM1-protein ligase 1 | 1.65 | 2E-25 | 3E-25 |
| 38 | Q9H257 | *CARD9* | Caspase recruitment domain-containing protein 9 | 1.62 | 7E-25 | 1E-24 |
| 39 | Q14643 | *ITPR1* | Inositol 1,4,5-trisphosphate receptor type 1 | 1.55 | 1E-24 | 2E-24 |
| 40 | P01817 | *IGHV2-5* | Immunoglobulin heavy variable 2-5 | 1.85 | 5E-23 | 7E-23 |
| 41 | Q6VAB6 | *KSR2* | Kinase suppressor of Ras 2 | 1.73 | 2E-22 | 3E-22 |
| 42 | Q6Y7W6 | *GIGYF2* | GRB10-interacting GYF protein 2 | 1.57 | 1E-21 | 1E-21 |
| 43 | Q9BQ69 | *MACROD1* | ADP-ribose glycohydrolase MACROD1 | 1.59 | 1E-21 | 2E-21 |
| 44 | P09619 | *PDGFRB* | Platelet-derived growth factor receptor beta | 1.69 | 1E-19 | 1E-19 |
| 45 | P20718 | *GZMH* | Granzyme H | 1.86 | 2E-16 | 2E-16 |
| 46 | O95696 | *BRD1* | Bromodomain-containing protein 1 | 1.62 | 7E-15 | 6E-15 |
| 47 | Q5SYB0 | *FRMPD1* | FERM and PDZ domain-containing protein 1 | 2.06 | 3E-14 | 2E-14 |
| 48 | O94851 | *MICAL2* | [F-actin]-monooxygenase MICAL2 | 1.64 | 9E-14 | 7E-14 |
| 49 | Q8WWB5 | *PIH1D2* | PIH1 domain-containing protein 2 | 1.63 | 9E-14 | 8E-14 |
| 50 | P01593 | *IGKV1D-33* | Immunoglobulin kappa variable 1D-33 | 1.86 | 1E-13 | 1E-13 |
| 51 | A0A087WSY6 | *IGKV3D-15* | Immunoglobulin kappa variable 3D-15 | 1.50 | 6E-12 | 4E-12 |
| 52 | Q7Z624 | *CAMKMT* | Calmodulin-lysine N-methyltransferase | 1.60 | 9E-12 | 6E-12 |
| 53 | P33316 | *DUT* | Deoxyuridine 5'-triphosphate nucleotidohydrolase, mitochondrial | 1.64 | 1E-10 | 7E-11 |
| 54 | Q8IYJ1 | *CPNE9* | Copine-9 | 1.54 | 2E-10 | 1E-10 |
| 55 | P00338 | *LDHA* | L-lactate dehydrogenase A chain | 1.89 | 2E-10 | 1E-10 |
| 56 | O60287 | *URB1* | Nucleolar pre-ribosomal-associated protein 1 | 1.72 | 2E-10 | 1E-10 |

| | | | | | | |
|---|---|---|---|---|---|---|
| 57 | P01715 | *IGLV3-1* | Immunoglobulin lambda variable 3-1 | 2.19 | 1E-09 | 7E-10 |
| 58 | Q6DN14 | *MCTP1* | Multiple C2 and transmembrane domain-containing protein 1 | 2.09 | 2E-09 | 1E-09 |
| 59 | A0A075B6I0 | *IGLV8-61* | Immunoglobulin lambda variable 8-61 | 1.53 | 3E-09 | 2E-09 |
| 60 | Q008S8 | *ECT2L* | Epithelial cell-transforming sequence 2 oncogene-like | 1.57 | 1E-08 | 6E-09 |
| 61 | P35398 | *RORA* | Nuclear receptor ROR-alpha | 1.50 | 2E-08 | 1E-08 |
| 62 | Q9NQU5 | *PAK6* | Serine/threonine-protein kinase PAK 6 | 1.56 | 5E-08 | 3E-08 |
| 63 | Q8IWY4 | *SCUBE1* | Signal peptide, CUB and EGF-like domain-containing protein 1 | 0.62 | 3E-07 | 2E-07 |
| 64 | O95477 | *ABCA1* | Phospholipid-transporting ATPase ABCA1 | 1.54 | 8E-07 | 4E-07 |
| 65 | P62805 | *H4C1* | Histone H4 | 2.88 | 6E-06 | 3E-06 |
| 66 | P24821 | *TNC* | Tenascin | 1.55 | 1E-05 | 6E-06 |
| 67 | Q9P2N5 | *RBM27* | RNA-binding protein 27 | 1.77 | 3E-05 | 1E-05 |
| 68 | A0A0J9YXX1 | *IGHV5-10-1* | Immunoglobulin heavy variable 5-10-1 | 0.66 | 5E-05 | 2E-05 |
| 69 | P45880 | *VDAC2* | Voltage-dependent anion-selective channel protein 2 | 0.63 | 2E-04 | 7E-05 |
| 70 | P07195 | *LDHB* | L-lactate dehydrogenase B chain | 2.09 | 2E-04 | 8E-05 |
| 71 | Q96D09 | *GPRASP2* | G-protein coupled receptor-associated sorting protein 2 | 1.94 | 2E-04 | 1E-04 |
| 72 | P00709 | *LALBA* | Alpha-lactalbumin | 1.59 | 3E-04 | 1E-04 |
| 73 | P01880 | *IGHD* | Immunoglobulin heavy constant delta | 1.55 | 5E-04 | 2E-04 |
| 74 | P00390 | *GSR* | Glutathione reductase, mitochondrial | 2.09 | 5E-04 | 2E-04 |
| 75 | A0A2R8Y619 | *H2BK1* | Histone H2B type 2-K1 | 2.44 | 7E-04 | 3E-04 |
| 76 | P0DJI8 | *SAA1* | Serum amyloid A-1 protein | 0.58 | 1E-03 | 4E-04 |
| 77 | Q86YZ3 | *HRNR* | Hornerin | 2.88 | 2E-03 | 6E-04 |
| 78 | Q96L34 | *MARK4* | MAP/microtubule affinity-regulating kinase 4 | 2.24 | 2E-03 | 8E-04 |
| 79 | O95803 | *NDST3* | Bifunctional heparan sulfate N-deacetylase/N-sulfotransferase 3; | 1.68 | 4E-03 | 1E-03 |
| 80 | Q8NCW5 | *NAXE* | NAD(P)H-hydrate epimerase | 1.73 | 4E-03 | 1E-03 |
| 81 | Q8TD10 | *MIPOL1* | Mirror-image polydactyly gene 1 protein | 1.75 | 8E-03 | 2E-03 |
| 82 | Q75N90 | *FBN3* | Fibrillin-3 | 0.49 | 9E-03 | 3E-03 |
| 83 | P18827 | *SDC1* | Syndecan-1 | 3.03 | 9E-03 | 3E-03 |
| 84 | P78371 | *CCT2* | T-complex protein 1 subunit beta | 1.68 | 1E-02 | 3E-03 |
| 85 | A0A0C4DH32 | *IGHV3-20* | Immunoglobulin heavy variable 3-20 | 0.56 | 1E-02 | 3E-03 |
| 86 | Q6NT04 | *TIGD7* | Tigger transposable element-derived protein 7 | 2.88 | 1E-02 | 3E-03 |
| 87 | O75110 | *ATP9A* | Probable phospholipid-transporting ATPase IIA | 1.64 | 2E-02 | 6E-03 |
| 88 | P11142 | *HSPA8* | Heat shock cognate 71 kDa protein | 1.60 | 3E-02 | 7E-03 |
| 89 | P10809 | *HSPD1* | 60 kDa heat shock protein, mitochondrial | 3.75 | 3E-02 | 9E-03 |
| 90 | P25786 | *PSMA1* | Proteasome subunit alpha type-1 | 6.22 | 5E-02 | 1E-02 |

[a]Fold change cutoff: Upregulation ≥1.5, Downregulation ≤0.66. Unpaired student's t-test with FDR correction was used for statistical evaluation in Spectronaut. [b] p-value <0.05 was considered statistically significant. [c] FDR-adjusted p-value (q-value) <0.05 was considered statistically significant. sPTB: Spontaneous preterm birth, TB: Term birth, EMT: Early-mid trimester.

**Table S3:** Protein-protein interaction subnetworks extracted from NetworkAnalyst.

| Subnetworks | EMT samples | | | LMT samples | | |
|---|---|---|---|---|---|---|
| | Nodes[a] | Edges[b] | Seeds[c] | Nodes[a] | Edges[b] | Seeds[c] |
| Subnetwork 1 | 1300 | 1550 | 58 | 1340 | 1587 | 41 |
| Subnetwork 2 | 59 | 68 | 2 | 54 | 59 | 2 |
| Subnetwork 3 | 17 | 16 | 2 | 13 | 12 | 1 |
| Subnetwork 4 | 13 | 12 | 1 | 10 | 9 | 1 |
| Subnetwork 5 | 10 | 9 | 1 | 8 | 7 | 1 |
| Subnetwork 6 | 8 | 7 | 1 | 8 | 7 | 1 |
| Subnetwork 7 | 8 | 7 | 1 | 7 | 6 | 1 |
| Subnetwork 8 | 8 | 7 | 1 | 6 | 5 | 1 |
| Subnetwork 9 | 8 | 7 | 1 | 5 | 4 | 1 |
| Subnetwork 10 | 7 | 6 | 1 | 5 | 4 | 1 |
| Subnetwork 11 | 6 | 5 | 2 | 5 | 4 | 1 |
| Subnetwork 12 | 5 | 4 | 1 | 5 | 4 | 1 |
| Subnetwork 13 | 5 | 4 | 1 | 4 | 3 | 1 |
| Subnetwork 14 | 5 | 4 | 1 | 4 | 3 | 1 |
| Subnetwork 15 | 4 | 3 | 1 | 4 | 3 | 1 |
| Subnetwork 16 | 4 | 3 | 1 | 3 | 2 | 1 |
| Subnetwork 17 | 4 | 3 | 1 | 3 | 2 | 1 |
| Subnetwork 18 | 3 | 2 | 1 | 3 | 2 | 1 |
| Subnetwork 19 | 3 | 2 | 1 | 3 | 2 | 1 |
| Subnetwork 20 | 3 | 2 | 1 | NA | NA | NA |

Subnetworks generated using the string interactome database with confidence score cutoff of 700. [a] Nodes represents the total no. of protein in the interactome including the seed proteins. [b]Edges denotes the total interactions between proteins in each sub-network. [c]Total query proteins in each sub-network.

**Table S4:** Peptides included in targeted proteomic assay (Phase 1- verification).

| Gene ID | Protein | Sequence | Annotation[a] | Transitions (precursor>product) |
|---|---|---|---|---|
| *ADGRA2* | Adhesion G Protein-Coupled Receptor A2 | LPFQC[160]SASYLGNDTR | ADGRA2_1C | 576.9391>505.237 |
| | | | ADGRA2_1D | 576.9391>891.4035 |
| | | NGSFLGLSLLEK | ADGRA2_4B | 639.3592>872.5457 |
| | | | ADGRA2_4D | 639.3592>389.24 |
| | | GGGALEKESHR | ADGRA2_5A | 570.791>243.109 |
| | | | ADGRA2_5B | 570.791>356.193 |
| *AVP* | Arginine Vasopressin | LVQLAGAPEPFEPAQPDAY | AVP_3A | 671.6709>465.1985 |
| | | | AVP_3C | 671.6709>213.1603 |
| | | GGKRAMSDLELR | AVP_4A | 444.907>545.787 |
| | | | AVP_4B | 444.907>243.145 |
| *CAPS2* | Calcyphosine 2 | AVISDPEQNLAIEQK | CAPS2_1A | 552.2936>588.3357 |
| | | | CAPS2_1B | 552.2936>517.2986 |
| | | AILQQGYADNSC[160]DK | CAPS2_4B | 528.2443>738.2728 |
| | | | CAPS2_4C | 528.2443>623.2459 |
| | | SQIQPFFGRK | CAPS2_7A | 604.333>311.171 |
| | | | CAPS2_7B | 604.333>329.182 |
| *CDC5L* | Cell Division Cycle 5 Like | ILLGGYQSR | CDC5L_5A | 503.788>667.316 |
| | | | CDC5L_5B | 503.788>780.4 |
| *CFTR* | CF Transmembrane Conductance Regulator | QAISPSDR | CFTR_5A | 437.225>561.263 |
| | | | CFTR_5B | 437.225>296.161 |
| *DPF1* | Double PHD Fingers 1 | GPGLAPGQIYTYPAR | DPF1_2D | 520.9444>678.3575 |
| | | ELAWVPEAQR | DPF1_3A | 599.8149>600.3105 |
| | | | DPF1_3C | 599.8149>885.4583 |
| | | SGHPSC[160]LQFTVNMTAAVR | DPF1_4C | 659.3213>558.2511 |
| *EGR4* | Early Growth Response 4 | MLHLSEFSEPDALLVK | EGR4_1B | 610.3225>378.2373 |
| | | | EGR4_1C | 610.3225>382.1913 |
| | | SDHLTTHVR | EGR4_4B | 355.8533>274.1879 |
| *FAM186A* | Family With Sequence Similarity 186 Member A | DTFAIESFR | FAM186A_3B | 543.2673>651.3466 |
| | | | FAM186A_3C | 543.2673>538.2625 |
| | | TSQISPLEWYQK | FAM186A_4C | 493.9213>438.2353 |
| | | KEEQVGEK | FAM186A_5A | 473.746>689.346 |
| | | | FAM186A_5C | 473.746>333.177 |
| *FRMPD1* | FERM And PDZ Domain Containing 1 | DVVYTYHQFIEAAK | FRMPD1_3A | 561.9513>735.3754 |
| | | | FRMPD1_3D | 561.9513>215.1032 |
| | | DIILTVK | FRMPD1_5B | 401.258>460.313 |
| *GIGYF2* | GRB10 Interacting GYF Protein 2 | NSNMGFWDDAVK | GIGYF2_3B | 692.3041>733.3521 |
| | | | GIGYF2_3D | 692.3041>880.4205 |
| | | KELEVQR | GIGYF2_5A | 451.259>531.289 |
| | | | GIGYF2_5B | 451.259>402.246 |
| *GRIPAP1* | Guanine nucleotide exchange factor for the Ras family of small G proteins | LQNSTLMAEFSK | GRIPAP1_1A | 684.8456>242.1505 |
| | | | GRIPAP1_1C | 684.8456>581.2935 |
| | | TNNYQLSDELR | GRIPAP1_5B | 451.553>532.273 |
| | | | GRIPAP1_5C | 451.553>417.246 |
| *GTF2IRD2* | GTF2I Repeat Domain Containing 2 | SGNEIFSR | GTF2IRD2_1B | 455.2254>409.2199 |
| | | FC[160]IDWSK | GTF2IRD2_4C | 478.2213>535.2516 |
| | | | GTF2IRD2_4D | 478.2213>308.1069 |
| | | IAELQTEFQK | GTF2IRD2_5A | 603.822>652.33 |
| | | | GTF2IRD2_5B | 603.822>422.24 |
| *GXYLT1* | Glucoside Xylosyltransferase 1 | GAGVAGPAAHPGVSDR | GXYLT1_3B | 709.8609>532.2682 |
| | | | GXYLT1_3D | 709.8609>767.38 |
| | | MHLAVVAC[160]GER | GXYLT1_4C | 414.8748>552.2968 |
| | | AVYEALR | GXYLT1_5A | 411.229>488.283 |
| | | | GXYLT1_5B | 411.229>651.346 |
| *IGKV1D-33* | Immunoglobulin Kappa Variable 1D-33 | DIQMTQSPSSLSASVGDR | IGKV1D-33_1A | 626.9675>691.3375 |
| | | | IGKV1D-33_1C | 626.9675>533.2683 |
| | | ILIYDASNLETGVPSR | IGKV1D-33_2A | 583.3129>616.3418 |
| | | | IGKV1D-33_2B | 583.3129>359.2043 |
| | | LLIYDASNLETGVPSR | IGKV1D-33_5A | 583.312>616.341 |
| | | | IGKV1D-33_5C | 583.312>515.294 |
| *IGLV2-8* | Immunoglobulin Lambda Variable 2-8 | VIIYEVNK | IGLV2-8_1C | 489.2875>765.4147 |
| | | | IGLV2-8_1D | 489.2875>261.1563 |
| | | LMIYEVSK | IGLV2-8_3A | 491.768>738.403 |
| | | | IGLV2-8_3B | 491.768>625.319 |
| *KIFC2* | Kinesin Family Member C2 | SLLALGGVMAALR | KIFC2_1B | 424.5889>561.3183 |
| | | | KIFC2_1C | 424.5889>359.2407 |
| | | VQHLTLENEALK | KIFC2_2A | 697.8861>584.3226 |
| | | | KIFC2_2C | 697.8861>816.4467 |

| | | | | |
|---|---|---|---|---|
| | | LQEAQDTTEALR | KIFC2_4B | 458.9007>589.3309 |
| *KRT25* | Keratin 25 | GLLSGNEK | KRT25_1A | 409.2249>647.3364 |
| | | | KRT25_1C | 409.2249>324.1721 |
| | | DAEAWFNEK | KRT25_2A | 555.2491>794.3837 |
| | | | KRT25_2C | 555.2491>723.3466 |
| | | VTMQNLNDR | KRT25_3A | 545.7697>631.3164 |
| | | | KRT25_3C | 545.7697>890.4154 |
| *KSR2* | Kinase Suppressor Of Ras 2 | NFNLPASHYYK | KSR2_1A | 677.3335>865.4208 |
| | | | KSR2_1D | 677.3335>473.24 |
| | | IVLDVNK | KSR2_5A | 400.748>588.335 |
| *LIN54* | Lin-54 DREAM MuvB Core Complex Component | LPFNGIIPSESASRPR | LIN54_1A | 580.9851>543.2904 |
| *MAP6* | Microtubule Associated Protein 6 | MVHETSYSAQFK | MAP6_1A | 476.5595>580.3095 |
| | | | MAP6_1C | 476.5595>294.1818 |
| | | DQDVVVPEHAK | MAP6_2A | 618.8151>581.3047 |
| | | | MAP6_2D | 618.8151>993.5369 |
| | | VKDQGSVVPESLKDQGPR | MAP6_6A | 647.013>714.378 |
| | | | MAP6_6B | 647.013>813.459 |
| *MDN1* | Midasin AAA ATPase 1 | GLSLGFLEK | MDN1_5A | 482.279>593.329 |
| | | | MDN1_5C | 482.279>536.308 |
| *MIA3* | Transport and Golgi organization protein 1 homolog | FGSTADALVSDDETTR | MIA3_1D | 562.2607>621.2844 |
| | | VQEENARLK | MIA3_5A | 543.799>859.463 |
| | | | MIA3_5B | 543.799>730.421 |
| *MICAL2* | [F-actin]-monooxygenase MICAL2 | GTLQAFNILTR | MICAL2_1B | 617.3517>763.4466 |
| | | LVLTQEQK | MICAL2_5A | 479.782>746.404 |
| | | | MICAL2_5C | 479.782>423.24 |
| *NAXE* | NAD(P)H-hydrate epimerase | GNAGGIQPDLLISLTAPK | NAXE5_2B | 589.0005>729.4511 |
| | | | NAXE5_2C | 589.0005>244.1661 |
| | | SGPTWWGPQR | NAXE5_3A | 586.2863>643.3316 |
| | | | NAXE5_3B | 586.2863>457.2523 |
| *PDGFRB* | Platelet-derived growth factor receptor beta | TLGDSSAGEIALSTR | PDGFRB_1A | 493.2552>547.3204 |
| | | | PDGFRB_1C | 493.2552>363.1992 |
| | | GDVALPVPYDHQR | PDGFRB_2B | 733.8735>506.2545 |
| | | | PDGFRB_2D | 733.8735>440.237 |
| | | DESVDYVPMLDMKGDVK | PDGFRB_5C | 647.638>848.918 |
| *PLEKHM1* | Pleckstrin homology domain-containing family M member 1 | EQPLESASDHPIASYR | PLEKHM1_1C | 600.624>706.3888 |
| | | ASSQDEAEDWLDR | PLEKHM1_4A | 761.3268>833.3794 |
| | | | PLEKHM1_4C | 761.3268>704.3368 |
| *PPAN* | Suppressor of SWI4 1 homolog | TEEELQAILEAK | PPAN_1A | 687.3621>772.4569 |
| | | | PPAN_1B | 687.3621>460.2771 |
| | | MTLQLIK | PPAN_4C | 423.7601>501.3401 |
| *RAB7A* | Ras-related protein Rab-7a | TSLMNQYVNK | RAB7A_1D | 599.3008>523.288 |
| | | ATIGADFLTK | RAB7A_4C | 518.7879>432.7455 |
| | | | RAB7A_4D | 518.7879>694.3776 |
| | | VIILGDSGVGK | RAB7A_5A | 529.316>845.473 |
| | | | RAB7A_5B | 529.316>732.389 |
| *RUNDC3A* | RUN domain-containing protein 3A | LQLQLEEAAAQNQR | RUNDC3A_1B | 806.4266>616.3167 |
| | | | RUNDC3A_1D | 806.4266>887.4335 |
| | | MSEYITTALR | RUNDC3A_2A | 592.8032>561.336 |
| | | | RUNDC3A_2D | 592.8032>460.2884 |
| | | GFWDYIR | RUNDC3A_5A | 478.735>752.373 |
| | | | RUNDC3A_5B | 478.735>566.293 |
| *SAA2* | Serum amyloid A-2 protein | SFFSFLGEAFDGAR | SAA2_1A | 775.8679>822.374 |
| | | | SAA2_1B | 775.8679>935.4581 |
| | | EANYIGSDK | SAA2_2A | 498.7358>406.1932 |
| | | | SAA2_2B | 498.7358>519.2773 |
| *SETD1B* | Histone-lysine N-methyltransferase SETD1B | YGEVEEVEILYNPK | SETD1B_1A | 841.4202>876.4831 |
| | | VDHDTIIDATK | SETD1B_3D | 409.8793>248.161 |
| | | VITVESQK | SETD1B_5A | 452.261>691.362 |
| | | | SETD1B_5B | 452.261>362.203 |
| *SPTY2D1* | Protein SPT2 homolog | GPGRPVSSPHELR | SPTY2D1_1D | 463.5852>277.6564 |
| | | PTVSSGPVPR | SPTY2D1_3A | 498.7778>699.3784 |
| | | | SPTY2D1_3D | 498.7778>525.3143 |
| | | RKEELVK | SPTY2D1_5A | 451.277>755.441 |
| | | | SPTY2D1_5B | 451.277>543.289 |
| *TC2N* | Tandem C2 domains nuclear protein | VGMFSSGELIYK | TC2N_3C | 665.8398>809.4409 |
| | | | TC2N_3D | 665.8398>665.3874 |
| | | IQTQTPR | TC2N_5A | 422.238>602.326 |
| | | | TC2N_5B | 422.238>356.696 |
| *TERF2* | Telomeric repeat-binding factor 2 | HEPGLGGPAER | TERF2_2B | 373.8568>472.252 |
| | | | TERF2_2D | 373.8568>529.2734 |

| | | | | |
|---|---|---|---|---|
| *TMC1* | Transmembrane channel-like protein 1 | FVSENEGALGK | TMC1_1A | 575.7911>904.4376 |
| | | VFTSWDYLIGNPETADNK | TMC1_2B | 690.6659>774.3634 |
| | | LKAELDEK | TMC1_5A | 473.266>391.182 |
| | | | TMC1_5B | 473.266>504.266 |
| *TMF1* | TATA element modulatory factor | MGAGSSIIENLQSQLK | TMF1_2B | 559.2948>603.3466 |
| | | LQVDMDELEEK | TMF1_3B | 674.8192>762.3521 |
| | | | TMF1_3C | 674.8192>518.2826 |
| *TOPAZ1* | Protein TOPAZ1 | NFSEVGFPDILK | TOPAZ1_1A | 683.3566>789.4511 |
| | | | TOPAZ1_1B | 683.3566>585.3612 |
| | | VITKEEK | TOPAZ1_5A | 423.75>634.341 |
| | | | TOPAZ1_5B | 423.75>533.293 |
| *UBE2O* | (E3-independent) E2 ubiquitin-conjugating enzyme | RPPEVFEQEIR | UBE2O_4B | 467.2497>674.3473 |
| | | | UBE2O_4C | 467.2497>288.2036 |
| | | FLDDIKK | UBE2O_5A | 439.753>618.346 |
| | | | UBE2O_5B | 439.753>731.43 |
| *VPS13B* | Vacuolar protein sorting-associated protein 13B | GPGAFVSGVSR | VPS13B_3B | 517.2754>505.2734 |
| | | | VPS13B_3C | 517.2754>604.3418 |
| | | LSEQQYNRLVDYITK | VPS13B_5B | 623.995>361.245 |
| *ZC3H10* | Zinc finger CCCH domain-containing protein 10 | HDLYDIYDLPDR | ZC3H10_3A | 512.2445>387.1992 |
| | | | ZC3H10_3B | 512.2445>615.3102 |
| | | KRVEELK | ZC3H10_5A | 451.277>642.357 |
| | | | ZC3H10_5B | 451.277>755.441 |

Peptides are presented with gene name followed by protein name and amino acid sequences. Transitions from precursor to product are shown. [a]Each transition is annotated with a unique code as follows: gene name_peptide number+transition alphabet.

**Table S5:** Predictive performance of Model 1 on test set at different prediction probability cutoffs.

| Classifiers | Threshold[a] | Test set (n=795) | | | | | | | | |
|---|---|---|---|---|---|---|---|---|---|---|
| | | Sensitivity | Specificity | PPV | NPV | Accuracy | TP | TN | FP | FN |
| SVM | 0.10 | 1.00 | 0.00 | 0.07 (0.05-0.08) | NA | 0.07 (0.05-0.08) | 52 | 0 | 743 | 0 |
| | 0.20 | 0.98 (0.93-1.0) | 0.00 | 0.06 (0.05-0.08) | 0.5 | 0.07 (0.05-0.08) | 51 | 1 | 742 | 1 |
| | 0.30 | 0.83 (0.72-0.92) | 0.2 (0.17-0.23) | 0.07 (0.05-0.09) | 0.94 (0.91-0.98) | 0.24 (0.21-0.27) | 43 | 147 | 596 | 9 |
| | 0.40 | 0.71 (0.58-0.83) | 0.43 (0.40-0.47) | 0.08 (0.06-0.11) | 0.96 (0.93-0.98) | 0.45 (0.41-0.48) | 37 | 321 | 422 | 15 |
| | 0.50 | 0.56 (0.42-0.69) | 0.53 (0.49-0.56) | 0.08 (0.05-0.10) | 0.94 (0.92-0.97) | 0.53 (0.49-0.56) | 29 | 391 | 352 | 23 |
| | 0.60 | 0.17 (0.08-0.29) | 0.84 (0.81-0.86) | 0.07 (0.03-0.12) | 0.94 (0.92-0.95) | 0.79 (0.77-0.82) | 9 | 622 | 121 | 43 |
| | 0.70 | 0.00 | 0.95 (0.93-0.96) | 0.00 | 0.93 (0.91-0.95) | 0.88 (0.86-0.91) | 0 | 703 | 40 | 52 |
| | 0.80 | 0.00 | 1.00 | NA | 0.93 (0.92-0.95) | 0.93 (0.92-0.95) | 0 | 743 | 0 | 52 |
| | 0.90 | 0.00 | 1.00 | NA | 0.93 (0.92-0.95) | 0.93 (0.92-0.95) | 0 | 743 | 0 | 52 |
| LR | 0.1 | 1.00 | 0.00 | 0.07 (0.05-0.08) | NA | 0.07 (0.05-0.08) | 52 | 0 | 743 | 0 |
| | 0.2 | 1.00 | 0.00 | 0.07 (0.05-0.08) | 1.00 | 0.07 (0.05-0.08) | 52 | 1 | 742 | 0 |
| | 0.3 | 0.98 (0.93-1.0) | 0.00 | 0.06 (0.05-0.08) | 0.67 | 0.07 (0.05-0.08) | 51 | 2 | 741 | 1 |
| | 0.4 | 0.94 (0.87-1.0) | 0.08 (0.06-0.10) | 0.07 (0.05-0.09) | 0.95 (0.89-1.0) | 0.14 (0.11-0.16) | 49 | 60 | 683 | 3 |
| | 0.5 | 0.13 (0.05-0.24) | 0.85 (0.82-0.88) | 0.06 (0.02-0.10) | 0.93 (0.91-0.95) | 0.8 (0.78-0.83) | 7 | 632 | 111 | 45 |
| | 0.6 | 0.00 | 1.00 | NA | 0.93 (0.92-0.95) | 0.93 (0.92-0.95) | 0 | 743 | 0 | 52 |
| | 0.7 | 0.00 | 1.00 | NA | 0.93 (0.92-0.95) | 0.93 (0.92-0.95) | 0 | 743 | 0 | 52 |
| | 0.8 | 0.00 | 1.00 | NA | 0.93 (0.92-0.95) | 0.93 (0.92-0.95) | 0 | 743 | 0 | 52 |
| | 0.9 | 0.00 | 1.00 | NA | 0.93 (0.92-0.95) | 0.93 (0.92-0.95) | 0 | 743 | 0 | 52 |
| RF | 0.1 | 1.00 | 0.00 | 0.07 (0.05-0.08) | 1.00 | 0.07 (0.05-0.09) | 52 | 1 | 742 | 0 |
| | 0.2 | 0.98 (0.94-1.0) | 0.05 (0.04-0.07) | 0.07 (0.05-0.09) | 0.97 (0.91-1.0) | 0.11 (0.09-0.13) | 51 | 38 | 705 | 1 |
| | 0.3 | 0.88 (0.80-0.96) | 0.24 (0.22-0.28) | 0.08 (0.05-0.10) | 0.97 (0.94-0.99) | 0.29 (0.26-0.32) | 46 | 182 | 561 | 6 |
| | 0.4 | 0.85 (0.74-0.94) | 0.32 (0.29-0.36) | 0.08 (0.06-0.10) | 0.97 (0.94-0.99) | 0.36 (0.32-0.39) | 44 | 241 | 502 | 8 |
| | 0.5 | 0.52 (0.38-0.65) | 0.5 (0.47-0.54) | 0.07 (0.04-0.09) | 0.94 (0.91-0.96) | 0.51 (0.47-0.54) | 27 | 375 | 368 | 25 |
| | 0.6 | 0.48 (0.34-0.61) | 0.58 (0.54-0.61) | 0.07 (0.05-0.10) | 0.94 (0.92-0.96) | 0.57 (0.54-0.61) | 25 | 429 | 314 | 27 |
| | 0.7 | 0.29 (0.17-0.42) | 0.73 (0.69-0.76) | 0.07 (0.04-0.11) | 0.94 (0.91-0.96) | 0.7 (0.67-0.73) | 15 | 540 | 203 | 37 |
| | 0.8 | 0.00 | 1.00 | NA | 0.93 (0.92-0.95) | 0.93 (0.92-0.95) | 0 | 743 | 0 | 52 |
| | 0.9 | 0.00 | 1.00 | NA | 0.93 (0.92-0.95) | 0.93 (0.92-0.95) | 0 | 743 | 0 | 52 |
| XGB | 0.1 | 1.00 | 0.00 | 0.07 (0.05-0.08) | NA | 0.07 (0.05-0.08) | 52 | 0 | 743 | 0 |
| | 0.2 | 1.00 | 0.00 | 0.07 (0.05-0.08) | NA | 0.07 (0.05-0.08) | 52 | 0 | 743 | 0 |
| | 0.3 | 1.00 | 0.00 | 0.07 (0.05-0.08) | NA | 0.07 (0.05-0.08) | 52 | 0 | 743 | 0 |
| | 0.4 | 0.94 (0.87-1.0) | 0.17 (0.15-0.20) | 0.07 (0.05-0.10) | 0.98 (0.95-1.0) | 0.22 (0.19-0.25) | 49 | 128 | 615 | 3 |
| | 0.5 | 0.48 (0.34-0.63) | 0.53 (0.49-0.56) | 0.07 (0.04-0.09) | 0.94 (0.91-0.96) | 0.53 (0.49-0.56) | 25 | 393 | 350 | 27 |
| | 0.6 | 0.12 (0.03-0.21) | 0.86 (0.84-0.89) | 0.06 (0.01-0.10) | 0.93 (0.91-0.95) | 0.82 (0.79-0.84) | 6 | 642 | 101 | 46 |
| | 0.7 | 0.00 | 1.00 | NA | 0.93 (0.92-0.95) | 0.93 (0.92-0.95) | 0 | 743 | 0 | 52 |

| | | | | | | | | | | |
|---|---|---|---|---|---|---|---|---|---|---|
| | 0.8 | 0.00 | 1.00 | NA | 0.93 (0.92-0.95) | 0.93 (0.92-0.95) | 0 | 743 | 0 | 52 |
| | 0.9 | 0.00 | 1.00 | NA | 0.93 (0.92-0.95) | 0.93 (0.92-0.95) | 0 | 743 | 0 | 52 |

AUROC: Area under the receiver operating curve, AURPC: Area under the precision recall curve. 95% CI was calculated by running 1000 iterations. Model was developed using the intensity of *MICAL2* and *CAPS2* in training set. PPV: Positive predictive value, NPV: Negative predictive value, TP: True positive, FP: False positive, TN: True negative, FN: False negative. [a] Prediction probability threshold.

**Table S6:** Predictive performance of Model 2 on test set at different prediction probability cutoffs.

| Classifiers | Threshold[a] | Test set (n=795) | | | | | | | | |
|---|---|---|---|---|---|---|---|---|---|---|
| | | Sensitivity | Specificity | PPV | NPV | Accuracy | TP | TN | FP | FN |
| SVM | 0.1 | 1.00 | 0.00 | 0.07 (0.05-0.08) | NA | 0.07 (0.05-0.08) | 52 | 0 | 743 | 0 |
| | 0.2 | 0.94 (0.87-1.0) | 0.15 (0.13-0.18) | 0.07 (0.05-0.09) | 0.97 (0.94-1.0) | 0.21 (0.18-0.23) | 49 | 114 | 629 | 3 |
| | 0.3 | 0.9 (0.82-0.98) | 0.18 (0.16-0.21) | 0.07 (0.05-0.09) | 0.96 (0.94-0.99) | 0.23 (0.20-0.26) | 47 | 137 | 606 | 5 |
| | 0.4 | 0.69 (0.57-0.82) | 0.42 (0.38-0.45) | 0.08 (0.05-0.10) | 0.95 (0.93-0.97) | 0.43 (0.40-0.47) | 36 | 309 | 434 | 16 |
| | 0.5 | 0.65 (0.52-0.78) | 0.44 (0.41-0.48) | 0.08 (0.05-0.10) | 0.95 (0.92-0.97) | 0.46 (0.42-0.49) | 34 | 330 | 413 | 18 |
| | 0.6 | 0.63 (0.50-0.76) | 0.45 (0.42-0.49) | 0.08 (0.05-0.10) | 0.95 (0.92-0.97) | 0.46 (0.43-0.50) | 33 | 336 | 407 | 19 |
| | 0.7 | 0.54 (0.41-0.68) | 0.65 (0.61-0.68) | 0.1 (0.06-0.13) | 0.95 (0.93-0.97) | 0.64 (0.60-0.67) | 28 | 480 | 263 | 24 |
| | 0.8 | 0.00 | 1.00 | NA | 0.93 (0.92-0.95) | 0.93 (0.92-0.95) | 0 | 743 | 0 | 52 |
| | 0.9 | 0.00 | 1.00 | NA | 0.93 (0.92-0.95) | 0.93 (0.92-0.95) | 0 | 743 | 0 | 52 |
| LR | 0.1 | 1.00 | 0.00 | 0.065 (0.05-0.08) | NA | 0.065 (0.05-0.08) | 52 | 0 | 743 | 0 |
| | 0.2 | 1.00 | 0.00 | 0.065 (0.05-0.08) | NA | 0.065 (0.05-0.08) | 52 | 0 | 743 | 0 |
| | 0.3 | 1.00 | 0.00 | 0.065 (0.05-0.08) | NA | 0.065 (0.05-0.08) | 52 | 0 | 743 | 0 |
| | 0.4 | 1.00 | 0.00 | 0.065 (0.05-0.08) | NA | 0.065 (0.05-0.08) | 52 | 0 | 743 | 0 |
| | 0.5 | 0.63 (0.50-0.76) | 0.45 (0.42-0.48) | 0.075 (0.05-0.10) | 0.946 (0.92-0.97) | 0.462 (0.43-0.50) | 33 | 334 | 409 | 19 |
| | 0.6 | 0.00 | 1.00 | NA | 0.935 (0.92-0.95) | 0.935 (0.91-0.95) | 0 | 743 | 0 | 52 |
| | 0.7 | 0.00 | 1.00 | NA | 0.935 (0.92-0.95) | 0.935 (0.92-0.95) | 0 | 743 | 0 | 52 |
| | 0.8 | 0.00 | 1.00 | NA | 0.935 (0.92-0.95) | 0.935 (0.92-0.95) | 0 | 743 | 0 | 52 |
| | 0.9 | 0.00 | 1.00 | NA | 0.935 (0.92-0.95) | 0.935 (0.91-0.95) | 0 | 743 | 0 | 52 |
| RF | 0.1 | 0.94 (0.87-1.0) | 0.16 (0.14-0.19) | 0.073 (0.05-0.09) | 0.98 (0.95-1.0) | 0.21 (0.19-0.24) | 49 | 121 | 622 | 3 |
| | 0.2 | 0.94 (0.88-1.0) | 0.17 (0.14-0.19) | 0.073 (0.05-0.09) | 0.98 (0.95-1.0) | 0.22 (0.19-0.25) | 49 | 124 | 619 | 3 |
| | 0.3 | 0.94 (0.88-1.0) | 0.17 (0.15-0.2) | 0.073 (0.06-0.09) | 0.98 (0.95-1.0) | 0.22 (0.19-0.25) | 49 | 127 | 616 | 3 |
| | 0.4 | 0.94 (0.87-1.0) | 0.17 (0.15-0.21) | 0.074 (0.06-0.09) | 0.98 (0.95-1.0) | 0.23 (0.19-0.25) | 49 | 130 | 613 | 3 |
| | 0.5 | 0.88 (0.79-0.97) | 0.23 (0.20-0.26) | 0.074 (0.05-0.10) | 0.97 (0.94-0.99) | 0.27 (0.24-0.31) | 46 | 171 | 572 | 6 |
| | 0.6 | 0.81 (0.69-0.91) | 0.42 (0.38-0.45) | 0.088 (0.06-0.11) | 0.97 (0.95-0.99) | 0.44 (0.41-0.48) | 42 | 311 | 432 | 10 |
| | 0.7 | 0.5 (0.36-0.64) | 0.69 (0.65-0.72) | 0.1 (0.06-0.14) | 0.95 (0.93-0.97) | 0.67 (0.64-0.70) | 26 | 509 | 234 | 26 |
| | 0.8 | 0.00 | 1.00 | NA | 0.93 (0.92-0.95) | 0.93 (0.92-0.95) | 0 | 743 | 0 | 52 |
| | 0.9 | 0.00 | 1.00 | NA | 0.93 (0.92-0.95) | 0.93 (0.92-0.95) | 0 | 743 | 0 | 52 |
| XGB | 0.1 | 0.942 (0.88-1.0) | 0.171 (0.14-0.20) | 0.074 (0.05-0.09) | 0.977 (0.95-1.0) | 0.221 (0.19-0.25) | 49 | 127 | 616 | 3 |
| | 0.2 | 0.923 (0.85-0.98) | 0.18 (0.15-0.21) | 0.073 (0.05-0.09) | 0.971 (0.94-0.99) | 0.229 (0.20-0.26) | 48 | 134 | 609 | 4 |
| | 0.3 | 0.923 (0.85-0.98) | 0.183 (0.16-0.21) | 0.073 (0.05-0.09) | 0.971 (0.94-0.99) | 0.231 (0.21-0.26) | 48 | 136 | 607 | 4 |
| | 0.4 | 0.904 (0.81-0.98) | 0.252 (0.22-0.28) | 0.078 (0.06-0.10) | 0.974 (0.95-0.99) | 0.294 (0.270.33) | 47 | 187 | 556 | 5 |
| | 0.5 | 0.865 (0.78-0.96) | 0.324 (0.29-0.36) | 0.082 (0.06-0.11) | 0.972 (0.94-0.99) | 0.36 (0.33-0.39) | 45 | 241 | 502 | 7 |
| | 0.6 | 0.865 (0.77-0.96) | 0.331 (0.30-0.37) | 0.083 (0.06-0.11) | 0.972 (0.94-0.99) | 0.366 (0.33-0.40) | 45 | 246 | 497 | 7 |
| | 0.7 | 0.827 | 0.393 | 0.087 | 0.972 | 0.421 | 43 | 292 | 451 | 9 |

| | | | | | | | | | | |
|---|---|---|---|---|---|---|---|---|---|---|
| | | (0.71-0.93) | (0.36-0.40) | (0.06-0.11) | (0.94-0.99) | (0.39-0.46) | | | | |
| | 0.8 | 0.346 (0.2-0.48) | 0.746 (0.71-0.77) | 0.087 (0.05-0.13) | 0.942 (0.92-0.96) | 0.719 (0.69-0.75) | 18 | 554 | 189 | 34 |
| | 0.9 | 0.000 | 1.000 | NA | 0.935 (0.92-0.95) | 0.935 (0.92-0.95) | 0 | 743 | 0 | 52 |

AUROC: Area under the receiver operating curve, AURPC: Area under the precision recall curve. 95% CI was calculated by running 1000 iterations. Model was developed using the intensity of *CAPS2* in training set. PPV: Positive predictive value, NPV: Negative predictive value, TP: True positive, FP: False positive, TN: True negative, FN: False negative. [a] Prediction probability threshold.

**Table S7:** Predictive performance of Model 3 on test set at different prediction probability cutoffs.

| Classifiers | Threshold[a] | Test set (n=795) | | | | | | | | |
|---|---|---|---|---|---|---|---|---|---|---|
| | | Sensitivity | Specificity | PPV | NPV | Accuracy | TP | TN | FP | FN |
| SVM | 0.1 | 1 | 0 | 0.065 (0.05-0.08) | NA | 0.065 (0.05-0.08) | 52 | 0 | 743 | 0 |
| | 0.2 | 1 | 0.004 (0-0.01) | 0.066 (0.05-0.08) | 1 | 0.069 (0.05-0.09) | 52 | 3 | 740 | 0 |
| | 0.3 | 0.942 (0.87-1) | 0.079 (0.06-0.10) | 0.067 (0.05-0.09) | 0.952 (0.89-1) | 0.136 (0.11-0.16) | 49 | 59 | 684 | 3 |
| | 0.4 | 0.846 (0.74-0.93) | 0.318 (0.28-0.35) | 0.08 (0.06-0.10) | 0.967 (0.95-0.99) | 0.352 (0.32-0.38) | 44 | 236 | 507 | 8 |
| | 0.5 | 0.481 (0.35-0.60) | 0.629 (0.59-0.66) | 0.083 (0.06-0.12) | 0.945 (0.92-0.97) | 0.619 (0.58-0.65) | 25 | 467 | 276 | 27 |
| | 0.6 | 0.154 (0.06-0.25) | 0.873 (0.85-0.90) | 0.078 (0.03-0.13) | 0.937 (0.92-0.95) | 0.826 (0.8-0.85) | 8 | 649 | 94 | 44 |
| | 0.7 | 0.019 (0-0.06) | 0.972 (0.96-0.98) | 0.045 (0-0.14) | 0.934 (0.92-0.95) | 0.909 (0.89-0.93) | 1 | 722 | 21 | 51 |
| | 0.8 | 0 | 1 | NA | 0.935 (0.92-0.95) | 0.935 (0.92-0.95) | 0 | 743 | 0 | 52 |
| | 0.9 | 0 | 1 | NA | 0.935 (0.92-0.95) | 0.935 (0.92-0.95) | 0 | 743 | 0 | 52 |
| LR | 0.1 | 0.942 (0.87-1) | 0.075 (0.06-0.09) | 0.067 (0.05-0.08) | 0.949 (0.88-1.0) | 0.132 (0.11-0.16) | 49 | 56 | 687 | 3 |
| | 0.2 | 0.808 (0.69-0.91) | 0.209 (0.18-0.24) | 0.067 (0.05-0.09) | 0.939 (0.9-0.97) | 0.248 (0.22-0.28) | 42 | 155 | 588 | 10 |
| | 0.3 | 0.712 (0.58-0.84) | 0.323 (0.29-0.36) | 0.069 (0.05-0.09) | 0.941 (0.91-0.97) | 0.348 (0.32-0.38) | 37 | 240 | 503 | 15 |
| | 0.4 | 0.635 (0.50-0.76) | 0.483 (0.45-0.52) | 0.079 (0.05-0.11) | 0.95 (0.93-0.97) | 0.493 (0.46-0.53) | 33 | 359 | 384 | 19 |
| | 0.5 | 0.462 (0.33-0.6) | 0.588 (0.55-0.63) | 0.073 (0.05-0.10) | 0.94 (0.92-0.96) | 0.58 (0.54-0.61) | 24 | 437 | 306 | 28 |
| | 0.6 | 0.346 (0.23-0.49) | 0.701 (0.67-0.74) | 0.075 (0.04-0.11) | 0.939 (0.92-0.96) | 0.678 (0.65-0.71) | 18 | 521 | 222 | 34 |
| | 0.7 | 0.212 (0.11-0.33) | 0.802 (0.77-0.83) | 0.07 (0.03-0.11) | 0.936 (0.92-0.95) | 0.764 (0.73-0.79) | 11 | 596 | 147 | 41 |
| | 0.8 | 0.077 (0.02-0.16) | 0.904 (0.88-0.93) | 0.053 (0.01-0.11) | 0.933 (0.91-0.95) | 0.85 (0.83-0.88) | 4 | 672 | 71 | 48 |
| | 0.9 | 0.038 (0-0.1) | 0.973 (0.96-0.98) | 0.091 (0-0.24) | 0.935 (0.92-0.95) | 0.912 (0.89-0.93) | 2 | 723 | 20 | 50 |
| RF | 0.1 | 0.981 (0.94-1) | 0.034 (0.02-0.05) | 0.066 (0.05-0.08) | 0.962 (0.88-1) | 0.096 (0.08-0.12) | 51 | 25 | 718 | 1 |
| | 0.2 | 0.923 (0.84-0.98) | 0.108 (0.09-0.13) | 0.068 (0.05-0.09) | 0.952 (0.9-0.99) | 0.161 (0.14-0.19) | 48 | 80 | 663 | 4 |
| | 0.3 | 0.808 (0.7-0.9) | 0.211 (0.18-0.24) | 0.067 (0.05-0.09) | 0.94 (0.9-0.97) | 0.25 (0.22-0.28) | 42 | 157 | 586 | 10 |
| | 0.4 | 0.654 (0.51-0.78) | 0.393 (0.36-0.43) | 0.07 (0.05-0.09) | 0.942 (0.91-0.96) | 0.41 (0.38-0.44) | 34 | 292 | 451 | 18 |
| | 0.5 | 0.481 (0.36-0.61) | 0.565 (0.53-0.60) | 0.072 (0.05-0.1) | 0.94 (0.91-0.96) | 0.56 (0.52-0.59) | 25 | 420 | 323 | 27 |
| | 0.6 | 0.288 (0.17-0.42) | 0.739 (0.71-0.77) | 0.072 (0.04-0.11) | 0.937 (0.92-0.96) | 0.709 (0.68-0.74) | 15 | 549 | 194 | 37 |
| | 0.7 | 0.154 (0.06-0.26) | 0.873 (0.85-0.9) | 0.078 (0.03-0.14) | 0.937 (0.92-0.95) | 0.826 (0.8-0.85) | 8 | 649 | 94 | 44 |
| | 0.8 | 0.019 (0-0.07) | 0.958 (0.94-0.97) | 0.031 (0-0.11) | 0.933 (0.91-0.95) | 0.897 (0.87-0.92) | 1 | 712 | 31 | 51 |
| | 0.9 | 0 | 1 | NA | 0.935 (0.92-0.95) | 0.935 (0.92-0.95) | 0 | 743 | 0 | 52 |
| XGB | 0.1 | 0.75 (0.63-0.86) | 0.276 (0.24-0.31) | 0.068 (0.05-0.09) | 0.94 (0.91-0.97) | 0.307 (0.27-0.34) | 39 | 205 | 538 | 13 |
| | 0.2 | 0.615 (0.48-0.74) | 0.377 (0.34-0.41) | 0.065 (0.04-0.09) | 0.933 (0.91-0.96) | 0.392 (0.36-0.43) | 32 | 280 | 463 | 20 |
| | 0.3 | 0.519 (0.39-0.66) | 0.443 (0.41-0.48) | 0.061 (0.04-0.08) | 0.929 (0.9-0.95) | 0.448 (0.41-0.48) | 27 | 329 | 414 | 25 |
| | 0.4 | 0.519 (0.39-0.66) | 0.503 (0.47-0.54) | 0.068 (0.04-0.09) | 0.937 (0.91-0.96) | 0.504 (0.47-0.54) | 27 | 374 | 369 | 25 |
| | 0.5 | 0.462 (0.33-0.6) | 0.575 (0.54-0.61) | 0.071 (0.05-0.1) | 0.938 (0.91-0.96) | 0.567 (0.53-0.6) | 24 | 427 | 316 | 28 |
| | 0.6 | 0.462 (0.33-0.6) | 0.614 (0.58-0.65) | 0.077 (0.05-0.11) | 0.942 (0.92-0.96) | 0.604 (0.57-0.64) | 24 | 456 | 287 | 28 |
| | 0.7 | 0.442 | 0.654 | 0.082 | 0.944 | 0.64 | 23 | 486 | 257 | 29 |

| | | | | | | | | | | |
|---|---|---|---|---|---|---|---|---|---|---|
| | | (0.30-0.59) | (0.62-0.69) | (0.05-0.12) | (0.91-0.96) | (0.61-0.68) | | | | |
| | 0.8 | 0.308 (0.19-0.43) | 0.725 (0.7-0.76) | 0.073 (0.04-0.11) | 0.937 (0.91-0.96) | 0.698 (0.67-0.73) | 16 | 539 | 204 | 36 |
| | 0.9 | 0.135 (0.05-0.24) | 0.808 (0.78-0.83) | 0.047 (0.02-0.08) | 0.93 (0.91-0.95) | 0.764 (0.73-0.79) | 7 | 600 | 143 | 45 |

AUROC: Area under the receiver operating curve, AURPC: Area under the precision recall curve. 95% CI was calculated by running 1000 iterations. Model was developed using RFE-CV selected socio-demographic and clinical parameters of participants in the training set. PPV: Positive predictive value, NPV: Negative predictive value, TP: True positive, FP: False positive, TN: True negative, FN: False negative.

[a] Prediction probability threshold.

**Table S8:** Predictive performance of Model 4 on test set at different prediction probability cutoffs.

| Classifiers | Threshold[a] | Test set (n=795) | | | | | | | | |
|---|---|---|---|---|---|---|---|---|---|---|
| | | Sensitivity | Specificity | PPV | NPV | Accuracy | TP | TN | FP | FN |
| SVM | 0.1 | 1 | 0.007 (0-0.01) | 0.066 (0.05-0.08) | 1 | 0.072 (0.06-0.09) | 52 | 5 | 738 | 0 |
| | 0.2 | 1 | 0.019 (0.01-0.03) | 0.067 (0.05-0.08) | 1 | 0.083 (0.06-0.1) | 52 | 14 | 729 | 0 |
| | 0.3 | 0.981 (0.93-1) | 0.044 (0.03-0.06) | 0.067 (0.05-0.09) | 0.971 (0.9-1) | 0.106 (0.09-0.13) | 51 | 33 | 710 | 1 |
| | 0.4 | 0.827 (0.72-0.93) | 0.18 (0.15-0.21) | 0.066 (0.05-0.08) | 0.937 (0.89-0.97) | 0.223 (0.19-0.25) | 43 | 134 | 609 | 9 |
| | 0.5 | 0.423 (0.29-0.57) | 0.611 (0.58-0.64) | 0.071 (0.04-0.1) | 0.938 (0.92-0.96) | 0.599 (0.56-0.63) | 22 | 454 | 289 | 30 |
| | 0.6 | 0.115 (0.04-0.22) | 0.929 (0.91-0.95) | 0.102 (0.03-0.18) | 0.938 (0.92-0.95) | 0.875 (0.85-0.9) | 6 | 690 | 53 | 46 |
| | 0.7 | 0 | 0.993 (0.99-1) | 0 | 0.934 (0.92-0.95) | 0.928 (0.91-0.95) | 0 | 738 | 5 | 52 |
| | 0.8 | 0 | 0.997 (0.99-1) | 0 | 0.934 (0.92-0.95) | 0.932 (0.91-0.95) | 0 | 741 | 2 | 52 |
| | 0.9 | 0 | 0.999 (0.99-1) | 0 | 0.935 (0.92-0.95) | 0.933 (0.92-0.95) | 0 | 742 | 1 | 52 |
| LR | 0.1 | 0.942 (0.88-1) | 0.078 (0.06-0.1) | 0.067 (0.05-0.09) | 0.951 (0.89-1) | 0.135 (0.11-0.16) | 49 | 58 | 685 | 3 |
| | 0.2 | 0.827 (0.72-0.92) | 0.195 (0.17-0.22) | 0.067 (0.05-0.09) | 0.942 (0.9-0.97) | 0.236 (0.21-0.27) | 43 | 145 | 598 | 9 |
| | 0.3 | 0.692 (0.56-0.81) | 0.332 (0.3-0.37) | 0.068 (0.05-0.09) | 0.939 (0.91-0.96) | 0.356 (0.32-0.39) | 36 | 247 | 496 | 16 |
| | 0.4 | 0.635 (0.51-0.76) | 0.482 (0.44-0.52) | 0.079 (0.05-0.11) | 0.95 (0.92-0.97) | 0.492 (0.46-0.53) | 33 | 358 | 385 | 19 |
| | 0.5 | 0.519 (0.37-0.65) | 0.592 (0.56-0.63) | 0.082 (0.05-0.11) | 0.946 (0.92-0.96) | 0.587 (0.55-0.62) | 27 | 440 | 303 | 25 |
| | 0.6 | 0.308 (0.2-0.45) | 0.693 (0.66-0.73) | 0.066 (0.04-0.1) | 0.935 (0.91-0.95) | 0.668 (0.64-0.7) | 16 | 515 | 228 | 36 |
| | 0.7 | 0.173 (0.07-0.29) | 0.818 (0.79-0.85) | 0.063 (0.03-0.11) | 0.934 (0.92-0.95) | 0.776 (0.75-0.8) | 9 | 608 | 135 | 43 |
| | 0.8 | 0.135 (0.05-0.23) | 0.907 (0.89-0.93) | 0.092 (0.04-0.16) | 0.937 (0.92-0.95) | 0.857 (0.83-0.88) | 7 | 674 | 69 | 45 |
| | 0.9 | 0.019 (0-0.07) | 0.972 (0.96-0.98) | 0.045 (0-0.15) | 0.934 (0.92-0.95) | 0.909 (0.89-0.93) | 1 | 722 | 21 | 51 |
| RF | 0.1 | 1 | 0.019 (0.01-0.03) | 0.067 (0.05-0.8) | 1 | 0.083 (0.07-0.1) | 52 | 14 | 729 | 0 |
| | 0.2 | 0.942 (0.87-1) | 0.092 (0.07-0.11) | 0.068 (0.05-0.09) | 0.958 (0.91-1) | 0.147 (0.12-0.17) | 49 | 68 | 675 | 3 |
| | 0.3 | 0.904 (0.81-0.98) | 0.211 (0.18-0.24) | 0.074 (0.05-0.09) | 0.969 (0.94-0.99) | 0.257 (0.23-0.29) | 47 | 157 | 586 | 5 |
| | 0.4 | 0.865 (0.77-0.95) | 0.31 (0.28-0.34) | 0.081 (0.06-0.11) | 0.97 (0.95-0.99) | 0.346 (0.31-0.38) | 45 | 230 | 513 | 7 |
| | 0.5 | 0.673 (0.55-0.8) | 0.451 (0.41-0.49) | 0.079 (0.06-0.11) | 0.952 (0.93-0.97) | 0.465 (0.43-5) | 35 | 335 | 408 | 17 |
| | 0.6 | 0.481 (0.34-0.62) | 0.641 (0.6-0.67) | 0.086 (0.05-0.12) | 0.946 (0.93-0.97) | 0.63 (0.6-0.66) | 25 | 476 | 267 | 27 |
| | 0.7 | 0.231 (0.12-0.35) | 0.806 (0.78-0.83) | 0.077 (0.04-0.12) | 0.937 (0.92-0.95) | 0.769 (0.74-0.8) | 12 | 599 | 144 | 40 |
| | 0.8 | 0.115 (0.03-0.21) | 0.914 (0.89-0.93) | 0.086 (0.03-0.15) | 0.937 (0.92-0.95) | 0.862 (0.84-0.88) | 6 | 679 | 64 | 46 |
| | 0.9 | 0 | 0.983 (0.97-0.99) | 0 | 0.934 (0.91-0.95) | 0.918 (0.9-0.94) | 0 | 730 | 13 | 52 |
| XGB | 0.1 | 1 | 0 | 0.065 (0.05-0.08) | NA | 0.065 (0.04-0.08) | 52 | 0 | 743 | 0 |
| | 0.2 | 0.962 (0.9-1) | 0.152 (0.13-0.18) | 0.074 (0.05-0.09) | 0.983 (0.95-1) | 0.205 (0.17-0.23) | 50 | 113 | 630 | 2 |
| | 0.3 | 0.923 (0.84-0.98) | 0.184 (0.16-0.22) | 0.073 (0.06-0.1) | 0.972 (0.94-0.99) | 0.233 (0.2-0.26) | 48 | 137 | 606 | 4 |
| | 0.4 | 0.885 (0.78-0.96) | 0.254 (0.22-0.28) | 0.077 (0.06-0.1) | 0.969 (0.94-0.99) | 0.296 (0.26-0.33) | 46 | 189 | 554 | 6 |
| | 0.5 | 0.635 (0.49-0.76) | 0.429 (0.39-0.47) | 0.072 (0.05-0.1) | 0.944 (0.92-0.97) | 0.443 (0.40-0.47) | 33 | 319 | 424 | 19 |
| | 0.6 | 0.538 (0.39-0.57) | 0.499 (0.46-0.53) | 0.07 (0.05-0.1) | 0.939 (0.92-0.96) | 0.502 (0.46-0.53) | 28 | 371 | 372 | 24 |
| | 0.7 | 0.212 (0.11-0.33) | 0.725 (0.69-0.76) | 0.051 (0.02-0.08) | 0.929 (0.91-0.95) | 0.692 (0.66-0.72) | 11 | 539 | 204 | 41 |

| | | | | | | | | | | |
|---|---|---|---|---|---|---|---|---|---|---|
| | 0.8 | 0.058 (0-0.13) | 0.915 (0.89-0.93) | 0.045 (0-0.1) | 0.933 (0.91-0.95) | 0.859 (0.83-0.88) | 3 | 680 | 63 | 49 |
| | 0.9 | 0 | 1 | NA | 0.935 (0.92-0.95) | 0.935 (0.91-0.95) | 0 | 743 | 0 | 52 |

AUROC: Area under the receiver operating curve, AURPC: Area under the precision recall curve. 95% CI was calculated by running 1000 iterations. Model was developed using RFE-CV selected socio-demographic and clinical parameters of participants along with *CAPS2* protein expression in the training set. PPV: Positive predictive value, NPV: Negative predictive value, TP: True positive, FP: False positive, TN: True negative, FN: False negative.
[a] Prediction probability threshold.

**Table S9:** Predictive performance of Model 2 on test set in high-risk sPTB cases at different prediction probability cutoffs.

| Classifiers | Threshold[a] | Test set (n=795) | | | | | | | | |
|---|---|---|---|---|---|---|---|---|---|---|
| | | Sensitivity | Specificity | PPV | NPV | Accuracy | TP | TN | FP | FN |
| SVM | 0.1 | 1 | 0 | 0.039 (0.03-0.05) | NA | 0.039 (0.03-0.05) | 31 | 0 | 764 | 0 |
| | 0.2 | 0.968 (0.88-1) | 0.152 (0.13-0.18) | 0.044 (0.03-0.06) | 0.991 (0.97-1) | 0.184 (0.16-0.21) | 30 | 116 | 648 | 1 |
| | 0.3 | 0.935 (0.84-1) | 0.183 (0.15-0.21) | 0.044 (0.03-0.06) | 0.986 (0.96-1) | 0.213 (0.19-0.24) | 29 | 140 | 624 | 2 |
| | 0.4 | 0.774 (0.6-0.91) | 0.416 (0.38-0.45) | 0.051 (0.03-0.07) | 0.978 (0.96-0.99) | 0.43 (0.39-0.46) | 24 | 318 | 446 | 7 |
| | 0.5 | 0.774 (0.62-0.92) | 0.446 (0.41-0.48) | 0.054 (0.03-0.08) | 0.98 (0.96-0.99) | 0.459 (0.42-0.49) | 24 | 341 | 423 | 7 |
| | 0.6 | 0.742 (0.57-0.9) | 0.454 (0.42-0.49) | 0.052 (0.03-0.07) | 0.977 (0.96-0.99) | 0.465 (0.43-0.5) | 23 | 347 | 417 | 8 |
| | 0.7 | 0.71 (0.55-0.86) | 0.648 (0.61-0.68) | 0.076 (0.05-0.11) | 0.982 (0.96-0.99) | 0.65 (0.62-0.68) | 22 | 495 | 269 | 9 |
| | 0.8 | 0 | 1 | NA | 0.961 (0.96-0.99) | 0.961 (0.95-0.97) | 0 | 764 | 0 | 31 |
| | 0.9 | 0 | 1 | NA | 0.961 (0.96-0.99) | 0.961 (0.95-0.97) | 0 | 764 | 0 | 31 |
| LR | 0.1 | 1 | 0 | 0.039 (0.03-0.05) | NA | 0.039 (0.03-0.05) | 31 | 0 | 764 | 0 |
| | 0.2 | 1 | 0 | 0.039 (0.03-0.05) | NA | 0.039 (0.03-0.05) | 31 | 0 | 764 | 0 |
| | 0.3 | 1 | 0 | 0.039 (0.03-0.05) | NA | 0.039 (0.03-0.05) | 31 | 0 | 764 | 0 |
| | 0.4 | 1 | 0 | 0.039 (0.03-0.05) | NA | 0.039 (0.03-0.05) | 31 | 0 | 764 | 0 |
| | 0.5 | 0.742 (0.58-0.89) | 0.452 (0.42-0.48) | 0.052 (0.03-0.07) | 0.977 (0.96-0.99) | 0.463 (0.43-0.5) | 23 | 345 | 419 | 8 |
| | 0.6 | 0 | 1 | NA | 0.961 (0.95-0.97) | 0.961 (0.95-0.97) | 0 | 764 | 0 | 31 |
| | 0.7 | 0 | 1 | NA | 0.961 (0.95-0.97) | 0.961 (0.95-0.97) | 0 | 764 | 0 | 31 |
| | 0.8 | 0 | 1 | NA | 0.961 (0.95-0.97) | 0.961 (0.95-0.97) | 0 | 764 | 0 | 31 |
| | 0.9 | 0 | 1 | NA | 0.961 (0.95-0.97) | 0.961 (0.95-0.97) | 0 | 764 | 0 | 31 |
| RF | 0.1 | 0.968 (0.89-1) | 0.161 (0.14-0.19) | 0.045 (0.03-0.06) | 0.992 (0.97-1) | 0.192 | 30 | 123 | 641 | 1 |
| | 0.2 | 0.968 (0.89-1) | 0.165 (0.14-0.19) | 0.045 (0.03-0.06) | 0.992 (0.97-1) | 0.196 | 30 | 126 | 638 | 1 |
| | 0.3 | 0.968 (0.89-1) | 0.169 (0.14-0.2) | 0.045 (0.03-0.06) | 0.992 (0.97-1) | 0.200 | 30 | 129 | 635 | 1 |
| | 0.4 | 0.968 (0.89-1) | 0.173 (0.15-0.2) | 0.045 (0.03-0.06) | 0.992 (0.97-1) | 0.204 | 30 | 132 | 632 | 1 |
| | 0.5 | 0.935 (0.84-1) | 0.229 (0.2-0.26) | 0.047 (0.03-0.06) | 0.989 (0.97-1) | 0.257 | 29 | 175 | 589 | 2 |
| | 0.6 | 0.871 (0.74-0.97) | 0.415 (0.38-0.45) | 0.057 (0.04-0.08) | 0.988 (0.97-1) | 0.433 | 27 | 317 | 447 | 4 |
| | 0.7 | 0.677 (0.5-0.83) | 0.687 (0.66-0.72) | 0.081 (0.05-0.12) | 0.981 (0.97-0.99) | 0.687 | 21 | 525 | 239 | 10 |
| | 0.8 | 0 | 1 | NA | 0.961 (0.95-0.97) | 0.961 | 0 | 764 | 0 | 31 |
| | 0.9 | 0 | 1 | NA | 0.961 (0.95-0.97) | 0.961 | 0 | 764 | 0 | 31 |
| XGB | 0.1 | 0.968 (0.89-1) | 0.169 (0.14-0.2) | 0.045 (0.03-0.06) | 0.992 (0.98-1) | 0.2 (0.17-0.23) | 30 | 129 | 635 | 1 |
| | 0.2 | 0.935 (0.83-1) | 0.178 (0.15-0.21) | 0.044 (0.03-0.06) | 0.986 (0.96-1) | 0.208 (0.18-0.24) | 29 | 136 | 628 | 2 |
| | 0.3 | 0.935 (0.83-1) | 0.181 (0.15-0.21) | 0.044 0.03-0.06) | 0.986 (0.96-1) | 0.21 (0.18-0.24) | 29 | 138 | 626 | 2 |
| | 0.4 | 0.935 (0.83-1) | 0.249 (0.22-0.28) | 0.048 0.03-0.06) | 0.99 (0.97-1) | 0.275 (0.24-0.31) | 29 | 190 | 574 | 2 |
| | 0.5 | 0.935 (0.83-1) | 0.322 (0.29-0.36) | 0.053 0.03-0.07) | 0.992 (0.98-1) | 0.346 (0.31-0.38) | 29 | 246 | 518 | 2 |
| | 0.6 | 0.935 (0.83-1) | 0.329 (0.3-0.36) | 0.054 0.04-0.07) | 0.992 (0.98-1) | 0.352 (0.32-0.39) | 29 | 251 | 513 | 2 |

| | | | | | | | | | | |
|---|---|---|---|---|---|---|---|---|---|---|
| | 0.7 | 0.903 (0.78-1) | 0.39 (0.36-0.42) | 0.057 (0.04-0.08) | 0.99 (0.98-1) | 0.41 (0.38-0.44) | 28 | 298 | 466 | 3 |
| | 0.8 | 0.387 (0.22-0.56) | 0.745 (0.71-0.78) | 0.058 (0.03-0.09) | 0.968 (0.95-0.98) | 0.731 (0.7-0.76) | 12 | 569 | 195 | 19 |
| | 0.9 | 0 | 1 | NA | 0.961 (0.95-0.97) | 0.931 (0.95-0.97) | 0 | 764 | 0 | 31 |

AUROC: Area under the receiver operating curve, AURPC: Area under the precision recall curve. 95% CI was calculated by running 1000 iterations. Model was developed using the intensity of CAPS2 in training set. It was tested on test set defining case samples at POG≤32. PPV: Positive predictive value, NPV: Negative predictive value, TP: True positive, FP: False positive, TN: True negative, FN: False negative. [a] Prediction probability threshold.
